# Trends in symptoms of anxiety and depression among adults in Norway: Evidence from eight population-based surveys (1995-2024)

**DOI:** 10.1101/2025.11.10.25339881

**Authors:** Benedicte Kirkøen, Astrid MA Eriksen, Ann Ragnhild Broderstad, Anne Høye, Jonas Johansson, Børge Sivertsen, Elin Skretting Lunde, Steinar Krokstad, Eivind Aakhus, Ellen Melbye Langballe, Thomas Sevenius Nilsen, Marit Knapstad, Thomas Hansen, Martin Tesli, Anne Reneflot

## Abstract

**Background:** Increasing symptoms of anxiety and depression among adolescents and emerging adults are well documented, while less attention has been given to how these symptoms have evolved among adults. We aim to describe the development of symptoms of anxiety and depression in the adult population in Norway over the past decades.

**Methods:** For the first time, we synthesize data from all population-based, cross-sectional surveys of symptoms of anxiety and depression among adults in Norway. Data were extracted from the Living Condition Survey (1998-2012, and 2015-2019), HUNT (1995/97-2017/19), the Tromsø Study (2001-2015/16), SAMINOR (2003/04–2012), Quality of Life Survey (2021-2024), the FHUS Agder (2019-2023), FHUS Oslo (2020-2024), and the Student’s Health and Wellbeing Study (SHoT, 2010-2022). Changes over time in symptoms were examined though sex and age stratified regression models.

**Results:** Data from the eight population-based studies, comprise 30 separate data collections including 584 173 adults aged 18-89 years between 1995 and 2024. We observe a clear increase in symptoms of anxiety and depression among young adults, especially among young women. Among individuals in midlife, findings are mixed, with most studies indicating stability or only minor fluctuations. In contrast, the symptoms of older adults appears to have remained stable or even improved.

**Conclusions:** Results show divergent trends in symptoms of anxiety and depression among adults. There has been an increase in symptoms among young adults, while symptoms remain stable or improved in old age. The results highlight the need for targeted preventive efforts among younger adults and for careful continued observation.

## Introduction

Mental health problems are among the most important contributors to disease burden worldwide [1]. For effective prevention and public health planning, it is crucial to have reliable estimates of changes in the prevalence of people living with mental disorders, as well as those experiencing high levels of symptoms that may develop into disorders.

Accumulating evidence indicates that symptoms of common mental disorders, such as anxiety and depression have increased rapidly over the past two decades, especially among girls and young women [2–7]. While many studies have examined trends in anxiety and depressive symptoms among adolescents, emerging adults, and university students [2–7], less attention has been given to how these symptoms have evolved among adults. Some studies have found an increase in anxiety and depressive symptoms (ADS) among adults over time [8–10] while others report stability [11]. Whereas some studies have reported trends for the entire adult population [11], an age-stratified study found significant increases in ADS among adults aged 20–29 and 30–39 years in Norway between 2006 and 2019 [6]. The increase was largest among women. In contrast, and during the same period, ADS remained stable or even improved among older adults. These findings underscore the importance of describing the trajectories of ADS that specific age-groups of men and women have had for the past decades.

A large study across 30 European countries showed substantial cross-national variation in trends in depressive symptoms [11]. This highlights that any meaningful assessment of whether ADS are increasing must start from a robust understanding of temporal developments within each country. Norway is fortunate to have several large population-based cross-sectional studies that track the health of the population through recurrent, self-reported measures. The studies cover different regions in Norway, one of which is designed specifically for the Indigenous population of Norway, the Sami, but also includes the rest of the population in the same geographic area [12, 13]. The studies include validated measures of ADS, but to date, only one study has published the development of ADS over time, showing trends up until 2019 [6]. The subsequent developments of ADS among adults in Norway remain unknown. For the first time, we will synthesize findings from all population-based surveys of self-reported ADS among adults in Norway, and provide a coherent picture of trends across different age-groups from 1995 until 2024. This allows us to make in-depth comparisons of ADS across different age groups, making it possible to identify specific age groups that may be more affected by changes in ADS over time. Moreover, combining data from several health surveys reduces the influence of study-specific factors, enhancing the validity of the findings. This approach will establish an important evidence base to inform policymaking and guide targeted interventions.

There are some challenges associated with comparing studies that measure ADS. The studies often use 1) different scales to measure symptoms of anxiety and depression. For instance, Hopkins Symptoms Checklist or Hospital Anxiety and Depression Scale. Or the studies use 2) different versions of the same scales (e.g., Hopkins Symptom Checklist have different versions; HSCL-25, HSCL-10 and HSCL-5), or 3) the same version of the same scale but with different cut-off levels to identify possible cases of anxiety and depression disorders from questionnaires. Further, results are frequently displayed for differing age-groups, further complicating comparisons. We aim to harmonize these data to make them as comparable as possible.

In the current study, we aim to synthesize data from population-based cross-sectional studies in Norway to describe the development of symptoms of anxiety and depression in the adult population over the past decades. By compiling and harmonizing data from multiple studies, we seek to provide a comprehensive and reliable overview of trends in ADS.

## Methods

### Inclusion criteria

All Norwegian repeated cross-sectional studies that measured symptoms of anxiety and depression in the general adult population (aged 18 +) were eligible for inclusion. For inclusion, each survey had to use a validated measure of symptoms of anxiety and depression, collect data for at least two time points, and recruit samples from the general population. In addition, one study recruiting university and college students in Norway was included. In Norway, 37.2% of individuals aged 19-24 years old enroll in higher education [14]. Thus, this group represents a distinct yet large segment of the adult population. Studies from clinical or at-risk populations were not included.

### Data Extraction

A systematic mapping of eligible studies was conducted by the research group at the Norwegian Institute of Public Health (NIPH). The preliminary list of studies was then shared with various research departments on mental health at NIPH, where researchers were encouraged to suggest other potential eligible studies. Once the overview was considered complete, principal investigators (PI) from all known eligible studies were contacted and invited to collaborate on the project in November 2022. The PIs were also asked if they were aware of other relevant studies that could be included. No additional studies were reported. This approach was necessary because not all population-based studies had published their results on trends of anxiety and depression symptoms in scientific journals. The data from each study were subsequently obtained either directly from the PI or from open repositories (data from the Living Condition Surveys (health and European health interview survey (EHIS)) [15–21] and the Quality of Life Surveys [22–26] were collected by Statistics Norway and obtained from the open repository sikt.no) between 2023-2025.

We extracted data on items measuring symptoms of anxiety and depression, participants’ age and sex as well as year of measurement. Additionally, the authors representing each survey provided study characteristics including sample size, measurement tools, the mode of measurement and response rate.

#### Measures

The included studies used one of the two validated measures of symptoms of anxiety and depression: The Hopkins Symptom Checklist (HSCL), or the Hospital Anxiety and Depression Scale (HADS).

### The Hopkins Symptom Checklist (HSCL)

The Hopkins symptoms checklist (HSCL) captures symptoms of anxiety and depression and asks the respondent to what extent different symptoms have bothered them for the past one or two weeks. Responses are scored on a Likert scale from 1 (not bothered’) to 4 (‘very bothered). The results are typically displayed as an average score or dichotomized based on established cut-off levels used to identify possible cases of mental disorders. Different studies use various short versions of the HSCL (HSCL-25, HSCL-10 and HSCL-5), and in order to harmonize data, the 5 items included in the HSCL-5 were extracted from larger scales where possible. In two studies (the Tromsø Study and SAMINOR), HSCL-10 was collected which includes only 4 of the 5 questions from HSCL-5, and therefore HSCL-10 results are displayed.

### HSCL-10

The short version HSCL-10 has been found to be a valid tool for measurement of symptoms of anxiety and depression in a Norwegian population [27], with the recommended cut-off point of ≥1.85. For participants lacking one or two items, missing values were replaced with the participant’s mean of the completed item scores. Data from participants with three or more missing items were excluded [28].

### HSCL-5

Two Norwegian studies showed that HSCL-5 is a valid tool for detection of anxiety and depression symptoms [27, 29]. A recent Spanish study proposed different cut-off levels for women (≥ 1.80) and men (≥ 2.00) [30], and the same cut-off levels were found to be most suitable in a Norwegian sample [29]. For the HSCL-5, the subscale of participants lacking more than two items was set to missing. For participants with one or two items missing, missing values were replaced with the mean of the completed item scores.

The HSCL-5 has also been validated specifically for the Norwegian student population [31]. That study showed that adopting slightly elevated cut-off values of ≥2.25 for male students and ≥2.75 for female students, enhanced the balance between sensitivity and specificity in this population.

### The Hospital Anxiety and Depression scale (HADS)

The Hospital Anxiety and Depression scale (HADS) consists of 14 questions, 7 measuring anxiety and 7 measuring depression symptoms [32]. Each question has the same four response options, ranging from 0–3, with a maximum total score of 21 for each scale. A higher score indicates higher levels of anxiety or depression. HADS has been validated in Norwegian background populations [33], with a score of ≥ 8 recommended as cut-off for possible caseness in HADS-anxiety and HADS-depression [33]. For participants lacking one or two items on one subscale, the missing values were substituted by the mean of the completed item scores. The subscale of participants lacking more than two items was set to missing.

#### Analyses

For each survey, we display the mean symptom scores and the percentage of individuals scoring above the pre-defined cut-off levels for being possible cases of mental disorders (percentage with mental health problems) across time, stratified by sex and 10-year age groups. To examine whether there were statistically significant changes over time in ADS within each survey, we conducted linear and logistic regression analyses. First, linear regression analyses were used to investigate changes in mean symptom scores over time for each survey separately. In these models, the mean symptom scores (dependent variable) were regressed on survey year and age (independent variables). Survey year was dummy-coded using a backward difference contrast coding scheme, allowing comparison between each survey year and the preceding one (i.e., 2002 vs. 1998; 2005 vs. 2002), and between the last and the first survey year. Analyses were stratified by sex and age-group.

Second, we performed logistic regression analyses to investigate changes over time in the proportion of individuals scoring above the cut-off levels. The analyses were stratified by sex and age group. Mental health problems were defined as scores above the cut-off levels ≥ 1.80 for women and ≥ 2.00 for men for HSCL-5 for general population [30], ≥2.75 for female and ≥2.25 for male students for HSCL-5 [31], ≥1.85 for HSCL-10 [27] and ≥8 for HADS-anxiety and HADS-depression [33].

One study (HUNT) employs HADS, and the range of possible scores in HADS differs from that of the other studies that employ HSCL (0-21 vs. 1-4). Therefore, the trend for mean symptom scores from the HUNT Study are shown in a separate figure. Additionally, while all other surveys provided data on the exact age of participants, SHoT only reported participants’ age groups (e.g. 18-20 years, 21-22 etc.) during its first data collections. Consequently, we could not align the data to the 20-29 age group used in the other studies. Instead, results are shown for the 18-28 age group in the student population. The age of invited participants differs between the studies. Consequently, we display data for studies where the age groups have at least two data collections with n>100 participants per data collection for each sex only. Owing to small sample sizes among participants aged 90+, data are presented for ages 18–89 years.

As can be seen in Table 1, the studies used various data collection methods, including paper questionnaire, telephone interviews, or web-based questionnaires. Since the mode of data collection is known to influence outcome measures [34–36], we display data collected by telephone separately. We defined the criteria for a clinically relevant change as a change of ½ standard deviation (s.d.), which is the smallest change perceived by individuals as an actual change in condition [37].

## Results

The study included eight repeated cross-sectional, population-based studies that measured symptoms of anxiety and depression in the general adult population in Norway (see Table 1). Together, they compromise 30 separate data collections, including 584 173 adults between 1995 and 2024. The age of participants ranged from 18 to 89 years, and between 49.8% and 54.4% % were women. Across surveys, older adults were more likely to participate than younger adults (Supplementary table S1), and the likelihood of participation increased with level of education. In the surveys that sampled participants across the country, there were few differences in participation between counties. See Table 1 for the characteristics of each survey. The surveys represent the general adult population, except for SHoT, which represents the overall student population. All studies, except the HUNT Study, used a version of HSCL to measure ADS. Here, we present results from analyses of changes from the first to last survey year for each study. Results from analyses comparing each survey year to the preceding one are found in the supplementary materials.

The included studies cover different age ranges, and therefore the set of studies represented differs across age groups. Absence of data from a study for a given age group indicates that no data were available for participants in that age range (e.g. The *SAMINOR Study* and the *Tromsø Study* does not include results for 20-29 age group).

### Trends in anxiety and depressive symptoms for women in different age groups

**For women 20-29 years**, the results show that ADS have increased across most studies. In more detail, there was a statistically significant increase in mean score (Figure 1a, 2a and supplementary material table 1-4a and 8-9a) and percentage scoring above cut-off (Figure 3a) from the first to the last measurement point for all surveys. An exception is the *FHUS Oslo Survey* that showed stability between 2020 and 2024. The changes observed for anxiety symptoms in HUNT were larger than ½ s.d. For students 18-28 years old, *the Student’s Health and Wellbeing Study (SHoT, 2010-2022)* showed a large and significant increase in mean score (Figure 1a, supplementary material table 5a) and percentage with mental health problems (Figure 3a).

**Fig. 1a.**
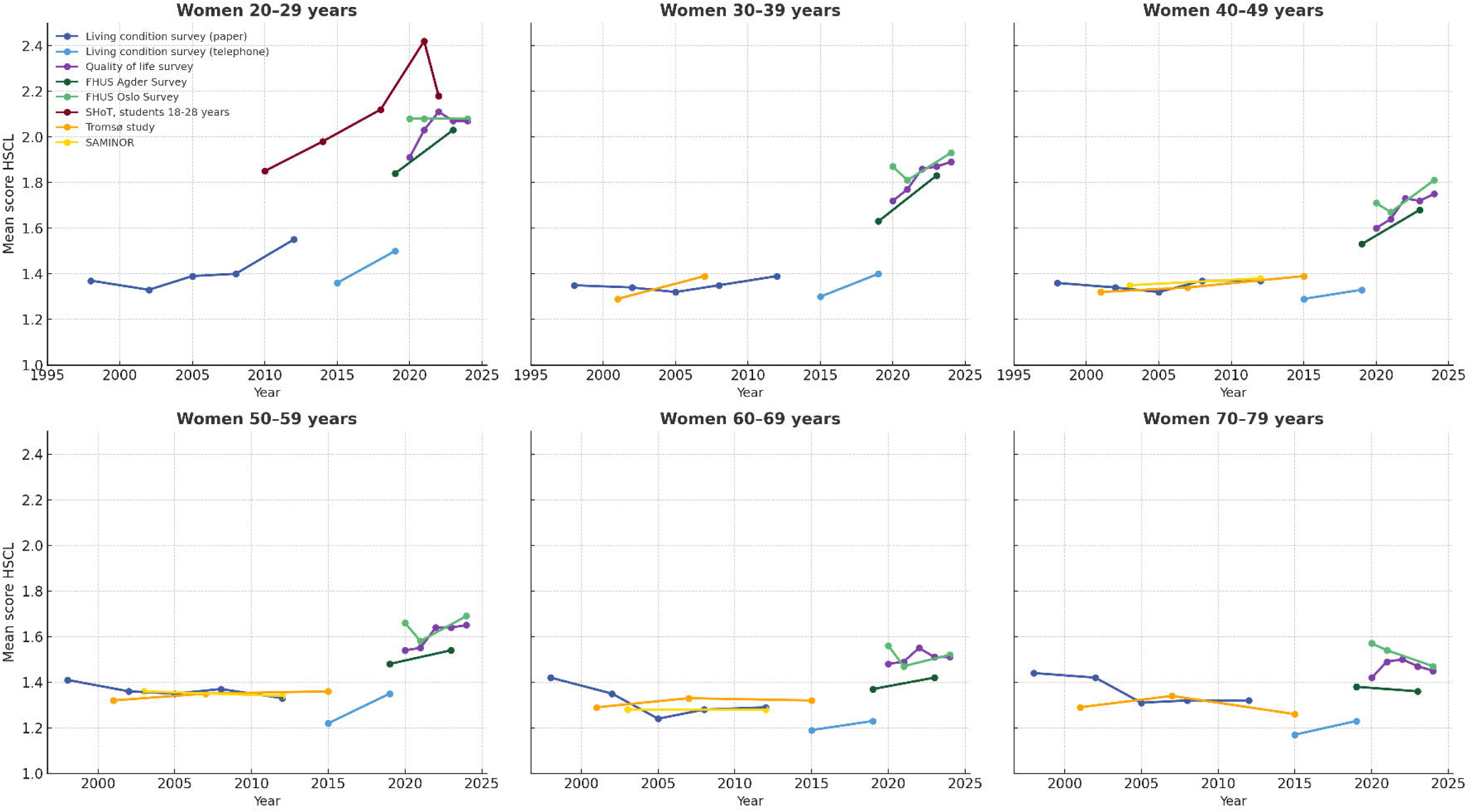
Trends for mean HSCL score for women in different age-groups Mean scores for Hopkins Symptom Checklist-5 (HSCL-5) are displayed for the Living Condition Survey (paper version and telephone version), the Quality of Life Survey, the FHUS Surveys (Agder and Oslo), and the SHoT study. Mean scores for HSCL-10 are displayed for the Tromsø Study and SAMINOR

**Fig. 1b.**
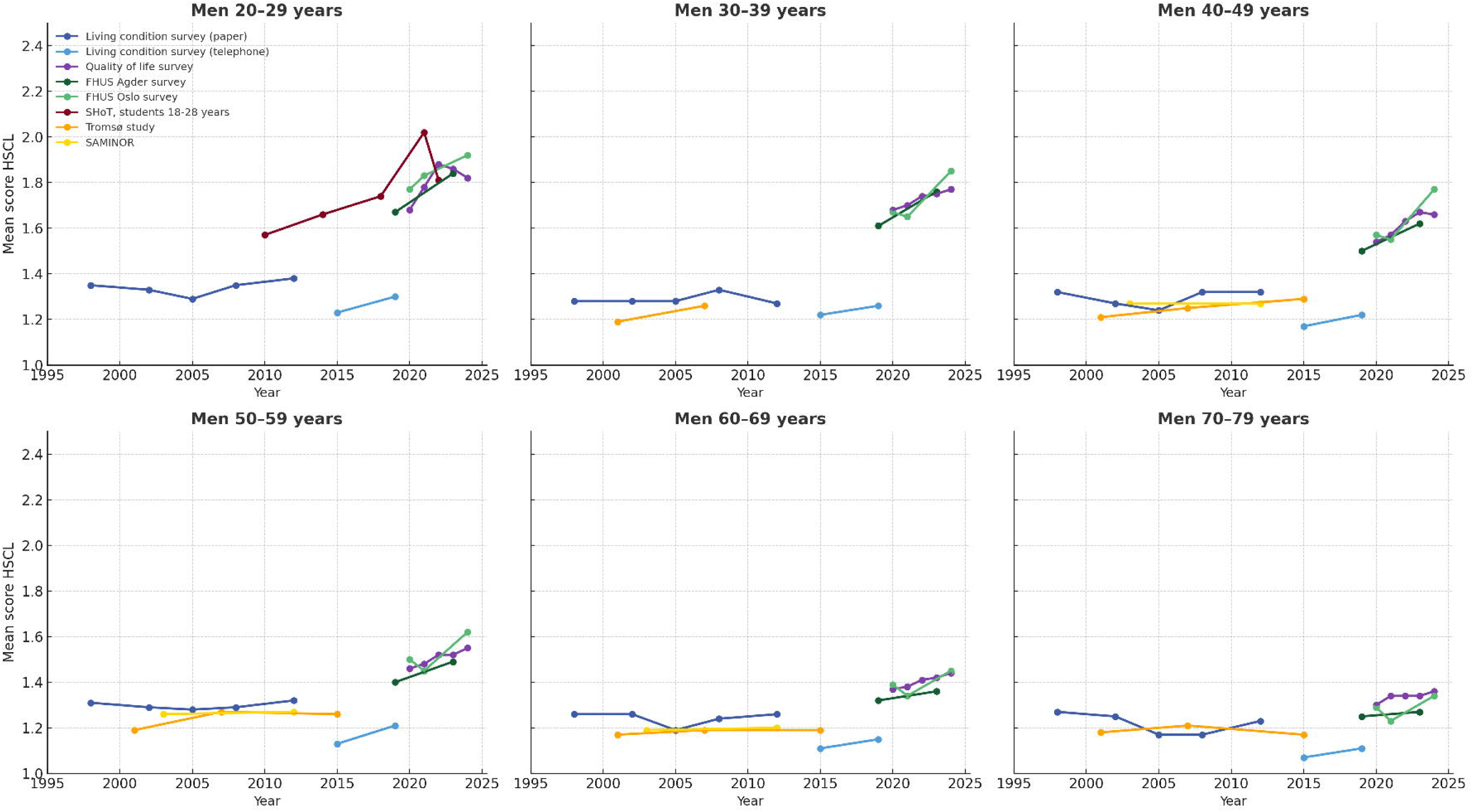
Trends for mean HSCL score for men in different age-groups Mean scores for Hopkins Symptom Checklist-5 (HSCL-5) are displayed for the Living Condition Survey (paper version and telephone version), the Quality of Life Survey, the FHUS Surveys (Agder and Oslo), and the SHoT study. Mean scores for HSCL-10 are displayed for the Tromsø Study and SAMINOR

**Fig. 1c.**
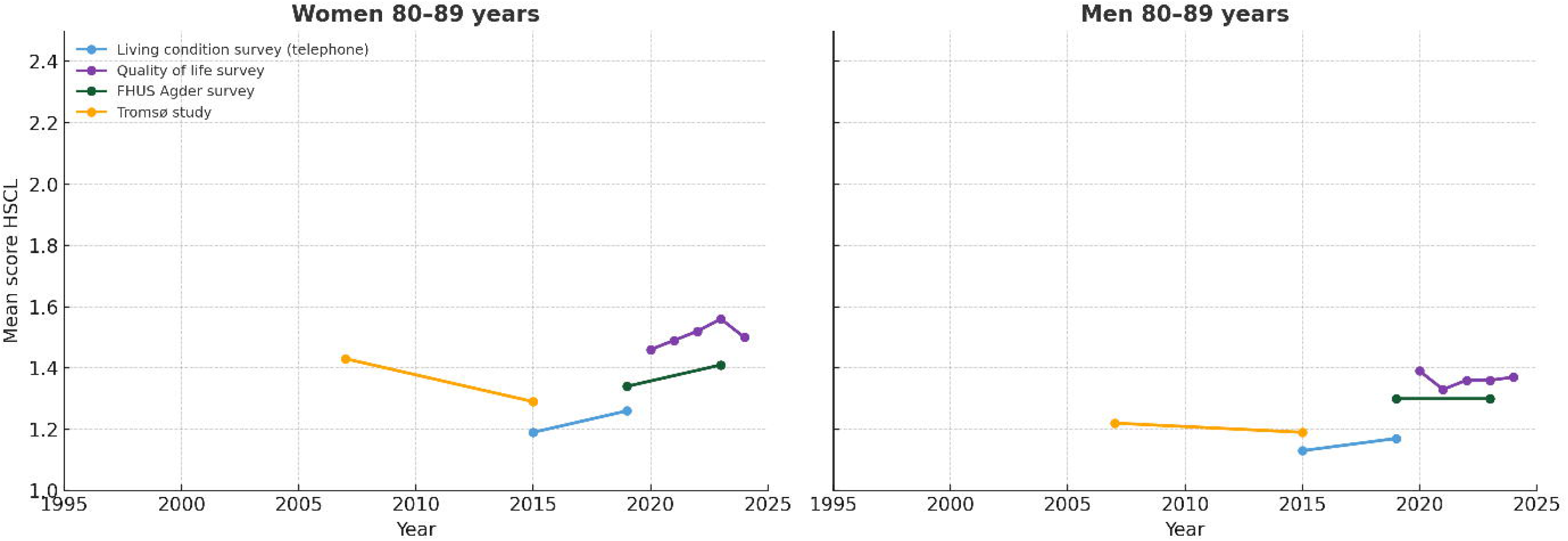
Trends for mean HSCL score for women and men 80-89 years old Mean scores for Hopkins Symptom Checklist-5 (HSCL-5) are displayed for the Living Condition Survey (telephone version), the Quality of Life Survey, the FHUS Agder Survey. Mean scores for HSCL-10 are displayed for the Tromsø Study. Studies with n<100 are not included.

**Fig. 2a.**
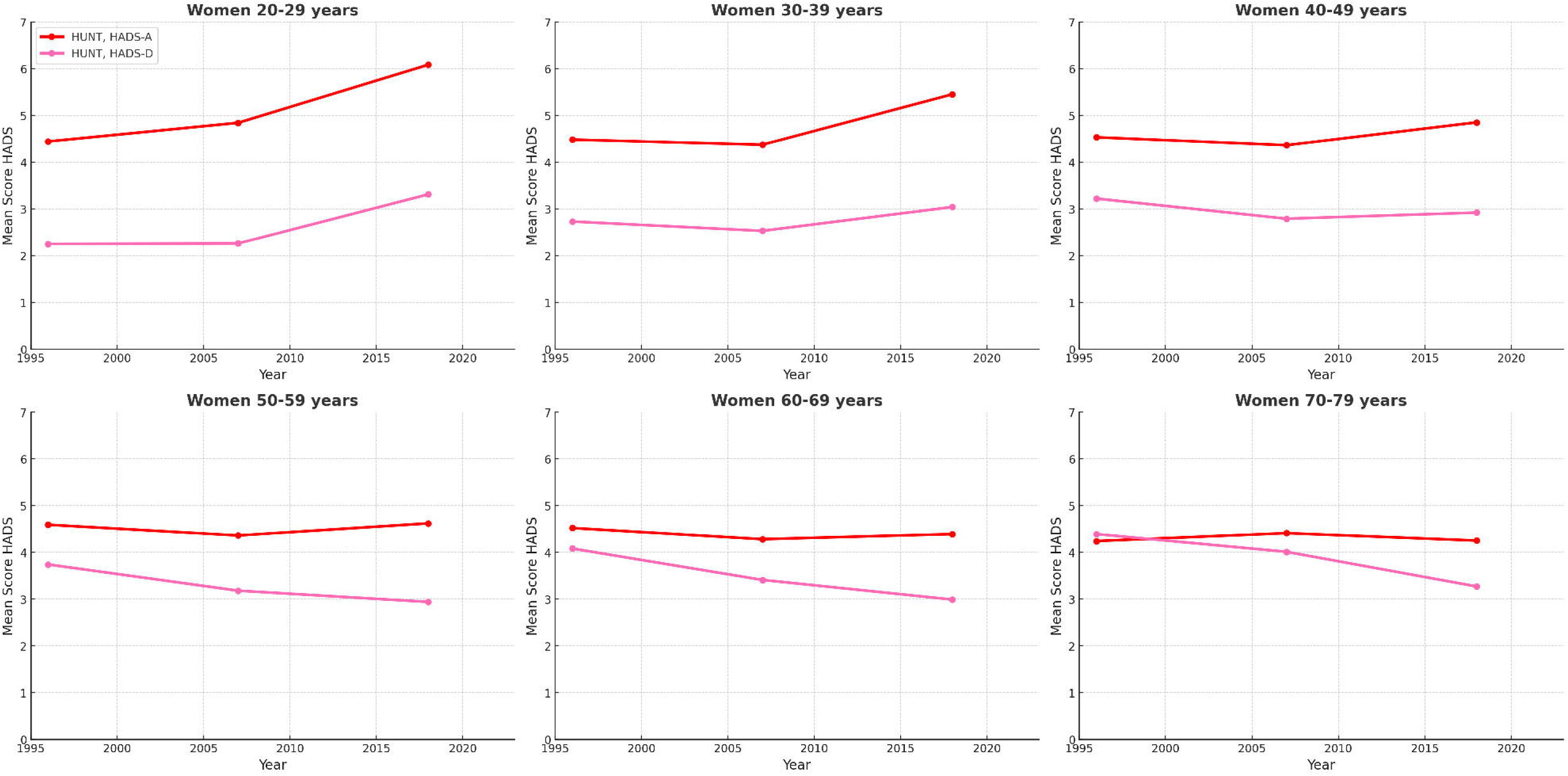
Trends for mean score on HADS-anxiety and HADS-depression for women in different age-groups

**Fig. 2b.**
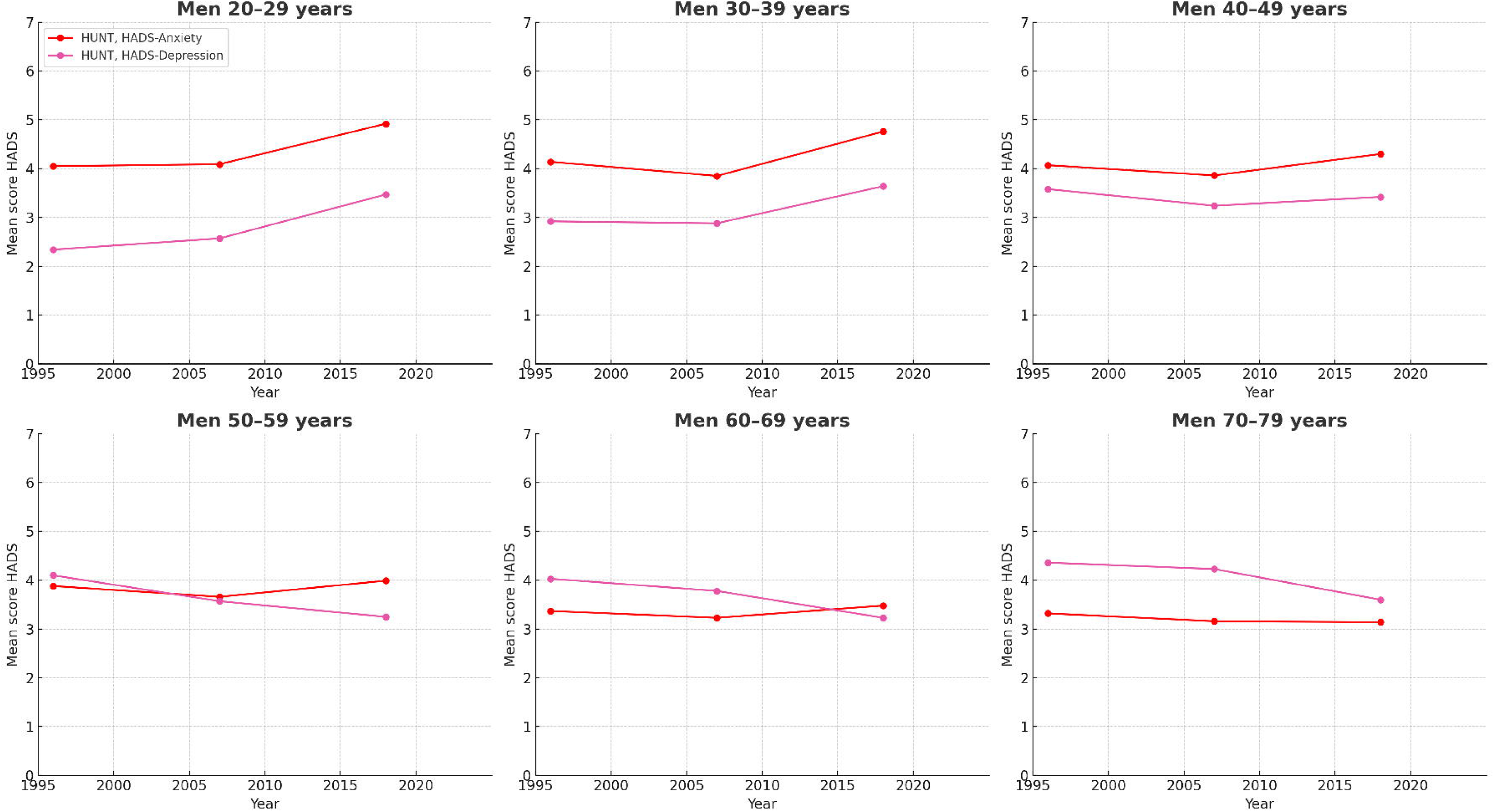
Trends for mean score on HADS-anxiety and HADS-depression for men in different age-groups

**Fig. 2c.**
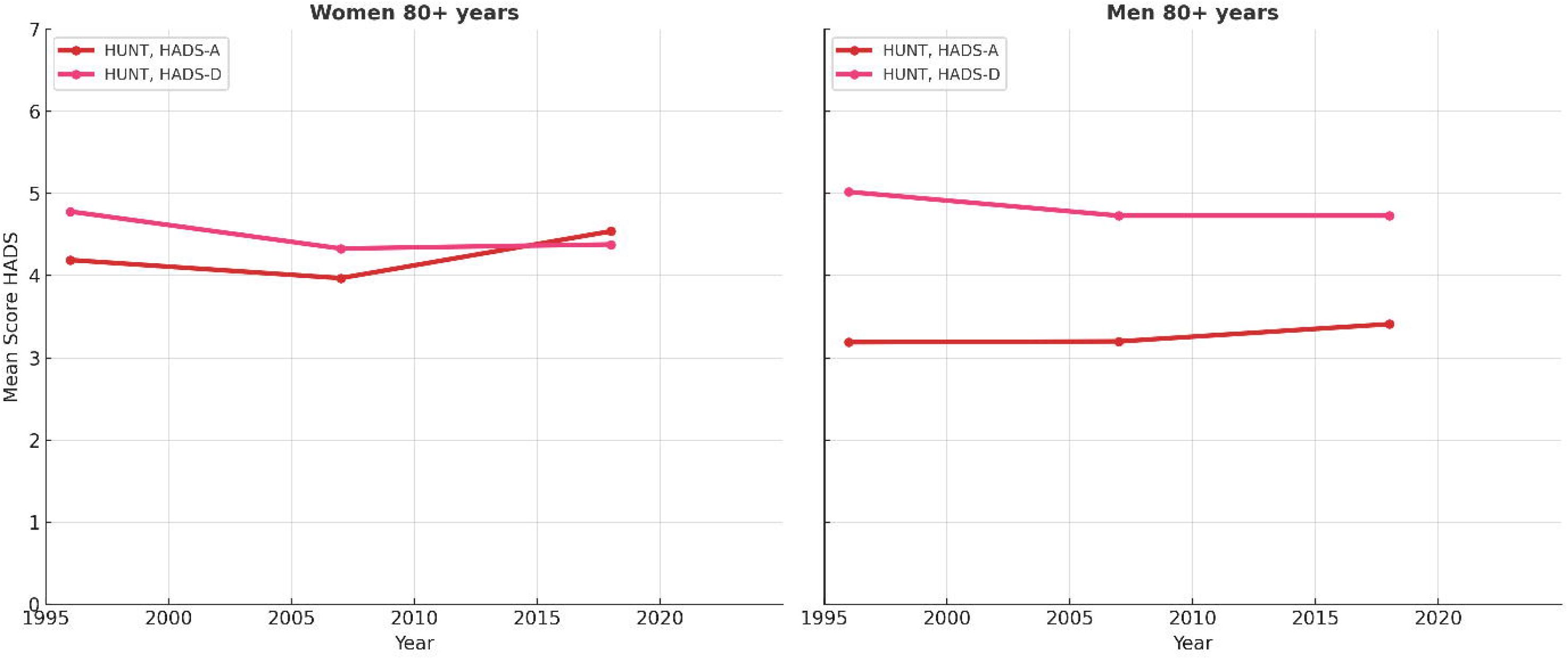
Trends for mean score on HADS-anxiety and HADS-depression for women and men 80-89 years old

**Fig. 3a.**
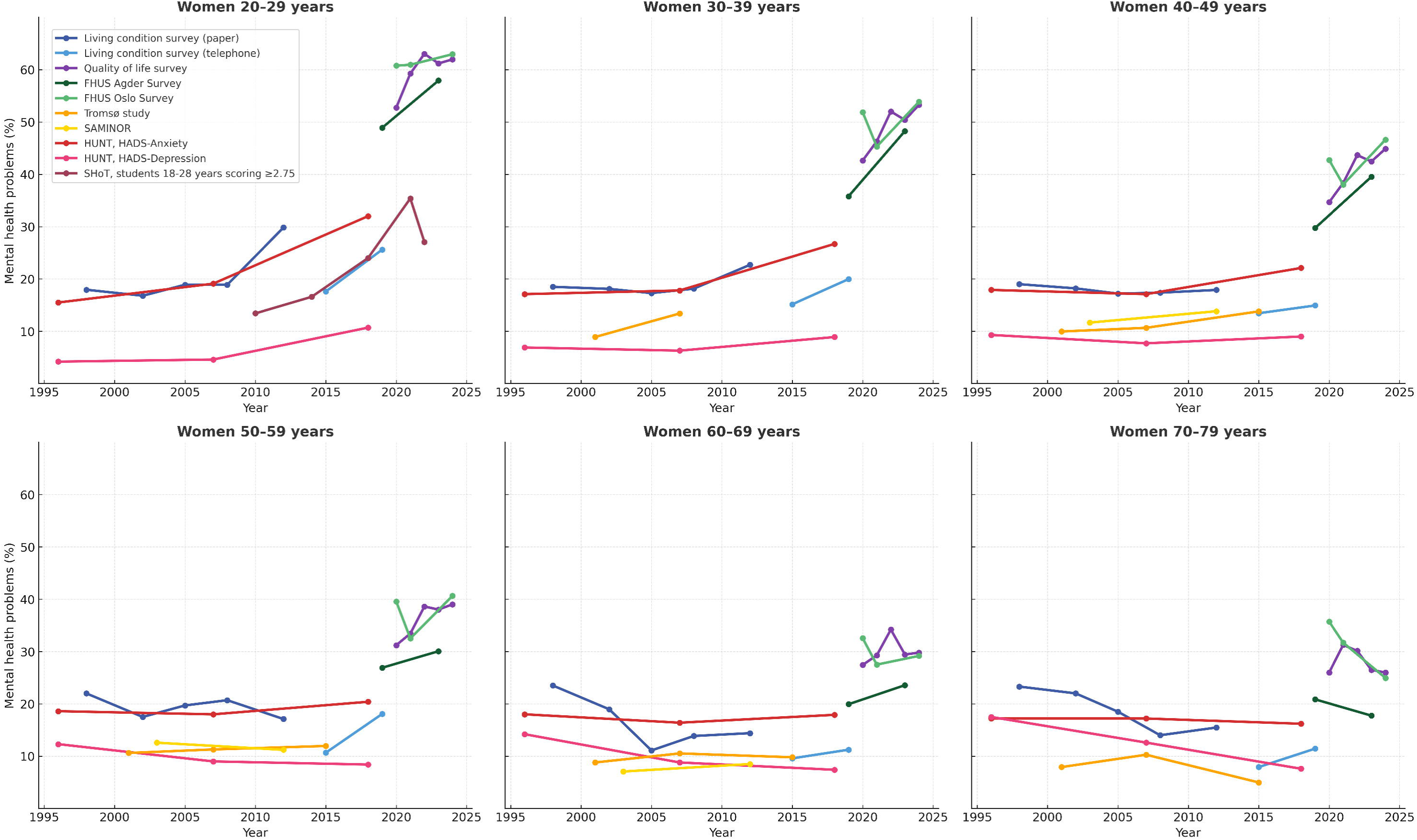
The percentage of women scoring above cut-off levels for mental health problems in different age-groups Mental health problems were defined as scores above the cut-off levels ≥ 1.80 for women for studies that use HSCL-5; Living Condition Survey (paper version and telephone version), the Quality of Life Survey, the FHUS surveys (Agder and Oslo). Mental health problems were defined as ≥2.75 for female students for HSCL-5. For the Tromsø Study and SAMINOR that use HSCL-10 mental health problems are defined as ≥1.85. In the HUNT Study mental health problems are defined as ≥8 for HADS-anxiety and HADS-depression.

**Fig. 3b.**
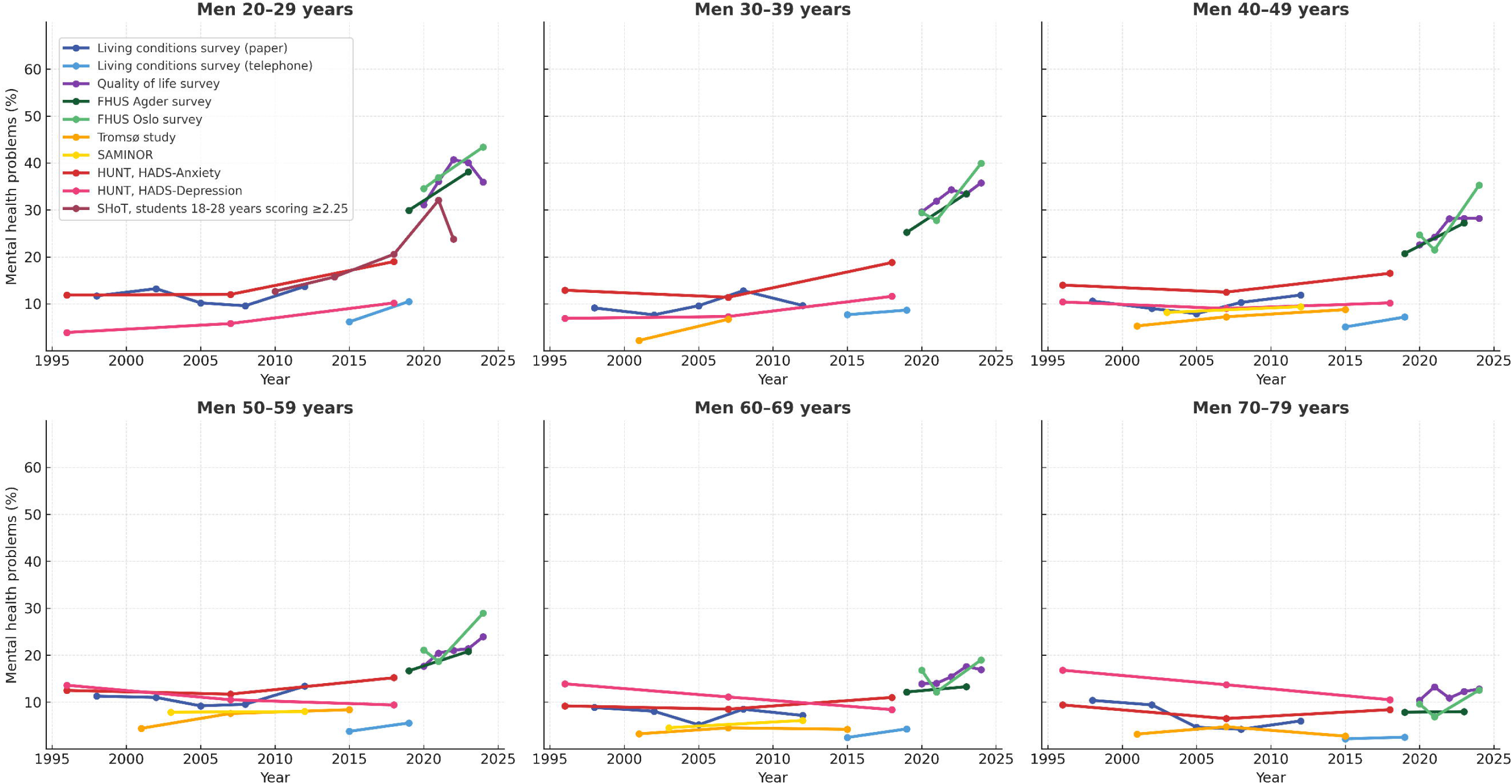
The percentage of men scoring above cut-off levels for mental health problems in different age-groups Mental health problems were defined as scores above the cut-off levels ≥ 2.00 for men for studies that use HSCL-5; Living Condition Survey (paper version and telephone version), the Quality of Life Survey, the FHUS Surveys (Agder and Oslo). Mental health problems were defined as ≥2.25 for male students for HSCL-5. For the Tromsø Study and SAMINOR that use HSCL-10 mental health problems are defined as ≥1.85. In the HUNT Study mental health problems are defined as ≥8 for HADS-anxiety and HADS-depression.

**Fig. 3c.**
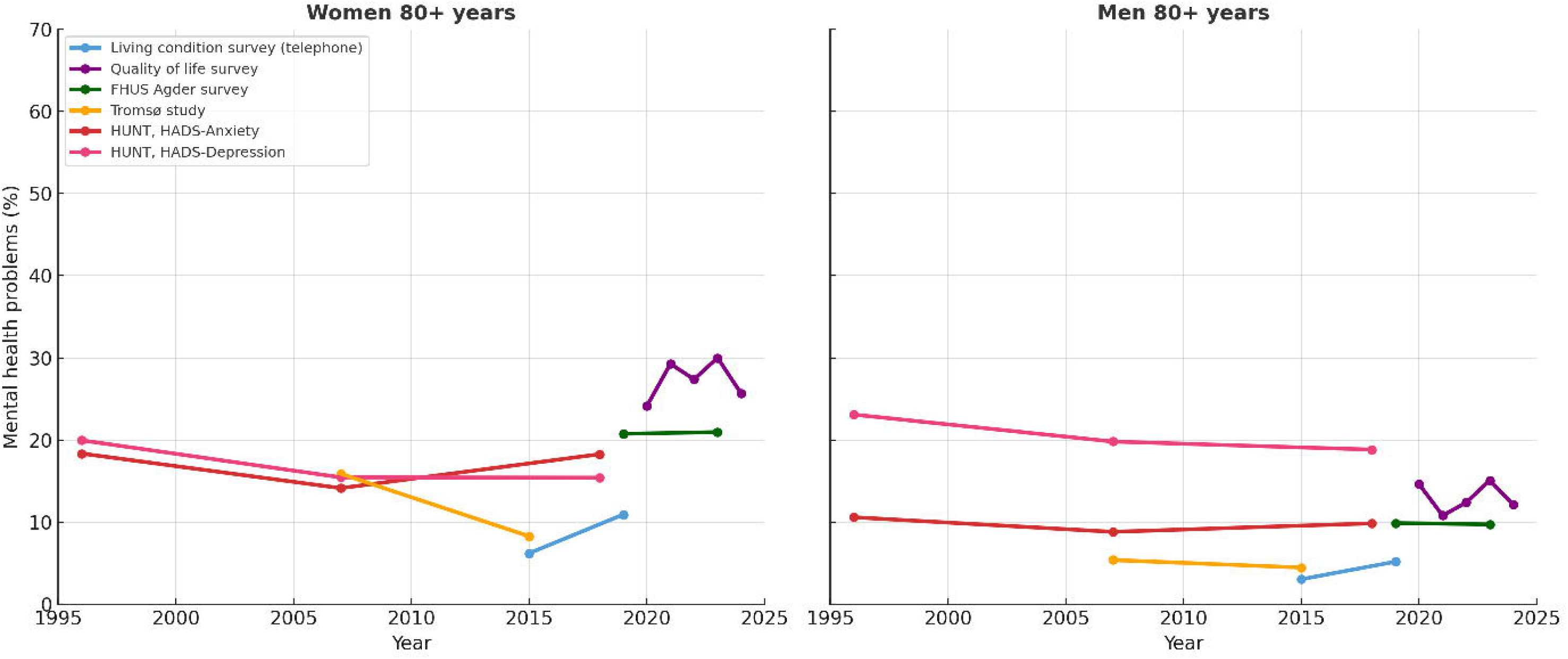
The percentage of women and men 80-89 years old scoring above cut-off levels for mental health problems Mental health problems were defined as scores above the cut-off levels ≥ 1.80 for women and ≥ 2.00 for men for studies that use HSCL-5; Living Condition Survey (telephone version), the Quality of Life Survey, the FHUS Agder survey. For the Tromsø Study that use HSCL-10 mental health problems are defined as ≥1.85. In the HUNT Study mental health problems are defined as ≥8 for HADS-anxiety and HADS-depression.

**Women, 30-39 years old.** In the *Living Condition Survey (paper, 1998-2012)* ADS remained stable. In the *Living Condition Survey (telephone version,2015-2019)*, *HUNT (1995/97-2017/19), the Tromsø Study (2001-2007/08), FHUS Agder (2019-2023) and the Quality of Life Survey (2021-2024)* there were significant increases in both the mean score and percentage with mental health problems (Figure 1-3a and supplementary material table 1-4a, 6a and 8-9a). *The FHUS Oslo Survey (2020-2024)* showed a significant increase in mean scores. Overall, findings suggest an increasing trend in ADS.

**Women, 40-49 years old.** Stability was observed in the *Living Condition Surveys (paper version 1998-2012 and telephone version 2015-2019)*. *HUNT (1995/97-2017/19)* showed that anxiety symptoms slightly increased over time, while depression mean score slightly decreased. The *Tromsø Study (2001–2015/16)* and *SAMINOR (2003/04–2012)* (Supplementary table 7a) showed slightly rising ADS, though the latter not consistently significant. While the Q*uality of Life Survey (2020–2024),* the *FHUS Surveys (Agder 2019-2023 and Oslo 2020-2024)* showed increased ADS. The results are mixed between stability and small increases in ADS, with a stronger increase in recent years.

However, symptoms of depression slightly decreased.

**Women, 50-59 years old.** Results across multiple surveys show mixed trends in ADS over time, with slight reductions in ADS in older datasets or in measures of depressive symptoms and increases in more recent ones. In the *Living Condition Survey (paper version, 1998–2012)*, ADS slightly decreased, while the following *Living Condition Survey (telephone version, 2015–2019)* indicates an increase. In the *HUNT Study (1995–2019),* a small but statistically significant increase in percentage with anxiety problems were observed, while depressive symptoms declined. The *Tromsø Study (2001–2015/16)* showed a minimal increase in mean score, while in the *SAMINOR Study (2003/04–2012)* and the *FHUS Oslo Survey (2020-2024)* ADS remained stable. The *FHUS Agder (2019-2023)* and the *Quality of Life Survey (2020–2024)* suggests an increase in ADS.

**Women, 60-69 years old.** In the *Living condition Survey (paper version, 1998–2012)*, ADS decreased significantly, followed by a minor increase in mean score in the *Living Condition Survey (telephone version, 2015–2019)*. The *HUNT Study (1995–2019)* indicated a decrease in depressive symptoms, while anxiety remained stable. The *FHUS Agder (2019-2023)* shows increases in ADS. The *Tromsø Study (2001–2015/16), SAMINOR (2003/04–2012), Quality of Life Survey (2020–2024)* and the *FHUS Oslo Survey (2020-2024)* showed no change. Overall, most studies suggest stable or decreasing ADS in this age group.

**Women, 70-79 years old.** Results show a general decline in ADS over time, with some fluctuation. In the *Living Condition Survey (paper version, 1998–2012),* ADS decreased. The *Living Condition Survey (telephone version, 2015–2019)* indicated a slight increase in mean score. The *HUNT Study (1995–2019)* found a reduction in depressive symptoms, while anxiety remained stable. The *Tromsø Study (2001–2015/16)* and the *FHUS Oslo Survey (2020-2024)* showed a decrease in ADS. In the *FHUS Agder (2019-2023)* and the *Quality of Life Survey (2021-2024)* ADS remained stable.

**Women, 80-89 years old.** The *HUNT Study (1995–2019)* showed a small increase in mean anxiety score only, while decreases were observed in depressive symptoms (Figure 1-3c). The *Tromsø Study (2007/08–2015/16)* showed a small decrease in ADS. There were no significant changes in the *Living Condition Survey (telephone version, 2015–2019), FHUS Agder (2019-2023)* or in the *Quality of Life Survey (2021-2024).* Overall, most studies suggest stable or decreasing ADS in this age group.

### Trends in anxiety and depressive symptoms for men in different age groups

**Men, 20-29 years old.** The *Living Condition Survey (paper version, 1998-2012)* showed no statistically significant changes (Figure 1b and 3b). However, in the *Living Condition Survey (telephone version, 2015-2019), HUNT (1995/97-2017/19), Quality of Life Survey (2020-2024) and the FHUS surveys (Agder 2019-2023, and Oslo 2020-2024)* there was an increase in mean scores (Figures 1-2b) and the proportion with mental health problems (Figure 3b, Supplementary material, Table 1-4b and 8-9b). For students aged 18-28 years, *SHoT (2010-2022)* showed a significant rise in ADS from 2010 to 2022 (Figure 1b and 3b, Supplementary material, Table 5b). Overall, multiple surveys indicate a significant rise in ADS over recent decades, though the increase is smaller compared to women in the same age group.

For **men aged 30-39 years**, ADS remained stable in the *Living Condition Survey (paper version, 1998-2012 and telephone version 2015-2019),* and the *Tromsø Study (2001-2007/08) (*supplementary material 6b). The *HUNT Study (1995/97-2017/19),* the *Quality of Life Survey (2020-2024)*, and *the FHUS Surveys (Agder 2019-2023 and Oslo 2020-2024)* showed statistically significant increases in ADS. In sum, early studies report stability, while recent studies report increases in ADS.

For **men aged 40-49 years**, no changes were observed in the *Living Condition Survey (paper version, 1998-2012) or the SAMINOR Study (2003/04-201,* Supplementary material, Table 7b). In the *Living Condition Survey (telephone version, 2015-2019)*, there was a small but statistically significant increase in the mean HSCL score only. Small but statistically significant increases in anxiety and decrease in depression mean scores were observed in the *HUNT Study (1995/97-2017/19). The Tromsø Study (2001-2015/16), the Quality of Life Survey (2020-2024) and the FHUS Surveys (Agder 2019-2023 and Oslo 2020-2024)* observed increases in ADS. Overall, early studies report a mix between stability and small increases in ADS and decreases in depressive symptoms. Recent studies report increases in ADS.

For **men aged 50-59 years**, findings across studies show mixed trends in ADS over time, with some studies finding no change and others observing small increases, while depressive symptoms decreased. No changes were observed in the *Living Condition Survey (paper version, 1998-2012)* or the *SAMINOR Study (2003/04-2012)*. *The Living Condition Survey (telephone version, 2015-2019)* showed a small increase in the mean HSCL score. In the *HUNT Study (1995/97-2017/19)*, there was a small increase in the percentage with anxiety problems, and a significant decrease depressive symptoms. Increases in ADS were observed in the *Tromsø Study (2001-2015/16),* the *Quality of Life Survey (2020-2024) and the FHUS Surveys (Agder 2019-2023 and Oslo 2020-2024)*.

For **men aged 60-69 years**, findings across studies indicate that there have been only small changes, with a decline in depressive symptoms. No changes were observed in the *Living Condition Survey (paper version, 1998-2012)* or the *SAMINOR Study (2003/04-2012)*. In the *Living Condition Survey (telephone version, 2015-2019),* the *Tromsø Study (2001-2015/16)* and t*he FHUS Surveys (Agder 2019-2023 and Oslo 2020-2024)* there were minor increases in mean HSCL scores. In the *HUNT Study (1995/97-2017/19)*, small but significant increases were observed for anxiety symptoms. For depressive symptoms, we observe a significant decrease. The *Quality of Life Survey (2020-2024)* showed small increases in ADS.

**Men aged 70-79 years.** No significant changes were observed in the *Living Condition Survey (paper, 1998-2012),* the *Tromsø Study (2001-2015/16) or the FHUS Surveys (Agder 2019-2023 and Oslo 2020-2024)*. The *Living Condition Survey (telephone, 2015-2019)* showed a small increase in the mean HSCL score. In the *HUNT Study (1995/97-2017/19)*, small decreases were observed in mean anxiety score as well as decreases in depressive symptoms. The *Quality of Life Survey (2020-2024)* reported a small increase in mean scores only. Overall, results indicate stability, or decreases in ADS, with some fluctuations.

For **men aged 80-89,** results indicate stability and decreases in depressive symptoms. There were no significant changes in the *Living Conditions Survey (telephone version, 2015–2019*), *the Tromsø Study (20007/08-2015/16), the FHUS Agder (2019-2023)* or in the *Quality of Life Survey (2021-2024)* (Figure 1-3c). *In* the *HUNT Study (1995/97-2017/19)* no changes were observed for anxiety symptoms, while the percentage of men scoring above cut off for depression decreased.

## Discussion

For the first time, utilizing Norway’s unique set of large population-based health surveys, we provide a comprehensive description of long-term trends in adult symptoms of anxiety and depression across sex and age groups. Overall, we observe a clear increase in ADS among young adults from 1995 to 2024, especially among young women. For adults in midlife, the results are mixed, with most studies indicating stability or only minor fluctuations, while the mental health of the old adults (up to age 89) appears to have improved or remained stable over the observation period. These divergent trends are in line with international studies [9, 38, 39].

Combining several studies enhances validity of the results, by reducing the influence of study-specific factors. To our knowledge, only two previous studies has examined adult ADS trends over time using multiple datasets, one focusing on three US studies from 1993–2020 [9] and one focusing on three studies of depressive symptoms in Germany between 2008-2023 [10]. The present study addresses four key limitations of these papers by relying exclusively on validated measures, reporting changes in symptoms of both anxiety and depression, harmonizing data to make studies comparable and extending the observation period from 1995 through 2024. Moreover, previous research [11] showed wide variability in trends between countries, indicating challenges with generalizing results across countries. Drawing on eight population-based studies, we provide a comprehensive and robust assessment of temporal changes in Norway.

The current paper points to a marked increase in ADS among young adults, particularly young women. Similar patterns have been reported in Sweden [40], Denmark [41], Scotland [39], Australia [38] and the US [2, 8, 9]. Our findings further demonstrate that the upward trend in mental health problems extends into young adulthood, up to the age of 39. Only a few international studies have reported similar patterns among adults in their thirties [2, 8, 38, 40], and the present study extends the observation period of these findings up to 2024. For the most recent time points, we also observe increased ADS in the 40-49 age group for both sexes, which should be closely monitored in the future. The observed increases were more pronounced among women. Sex differences in mental health problems are among the most stable findings in psychiatry [42], though the underlying causes remain unclear. However, women are exposed to many known risk factors.

Our results show stability in ADS among older adults, consistent with international studies reporting stability in anxiety symptoms in this age group [2]. For depressive symptoms, we observe a reduction, which aligns with findings from international studies [43–46]. The observed reduction in depressive symptoms coincides with a decline in loneliness among older adults [47]. While our study display an uplifting trend for mental health in old age, we should note that a more detailed analysis of older adults in HUNT shows decreases in depressive symptoms over time, with the exception of adults above 85 years old [48].

A key issue is whether the observed changes are large enough to be considered meaningful. The increase in symptoms of anxiety among woman aged 20-29 in HUNT exceeded ½ s.d., indicating a clinically relevant change [37]. The increase in ADS among female students, and in depressive symptoms among young men and women in HUNT were also close to a change of ½ s.d. In a previous study we showed that 21% of individuals scoring above the HSCL-5 cut-off met the diagnostic criteria for anxiety or depression disorders in a structured diagnostic interview [29]. Assuming this relation has remained stable over time, an estimated 3.8% of women aged 20–29 years met the criteria in 1998 (21% of the 18% above the cut-off in the Living Conditions Survey), compared to 13.0% in 2024 (21% of 62%). This change indicates a substantial public health impact.

Another core question is whether the observed changes are due to age, period or cohort effects. In the present study, age was held constant, yet trends changed, suggesting that patterns in young age groups cannot be explained by a pure age-effect. Period effects imply changes across all individuals regardless of age or cohort. Our data suggest stability in ADS until 2010, followed by increases among younger groups, indicating that period effects are not independent of age. The 2019/2020 surveys (Quality of Life Survey and FHUS) show rising ADS that parallel pandemic restrictions in Norway between 2020–2022. For women, increases appeared across all age groups, consistent with a period effect. While levels returned to 2020 levels among older adults after restrictions were lifted in 2022, they remained elevated among young adults. Cohort effects were not examined. Future research should clarify whether the observed trends reflect age–period interactions or true cohort effects.

The causes of increasing ADS among young adults remain unknown. Possible explanations include changes in reporting behavior, population composition, or societal conditions affecting mental health. Some argue that reduced stigma and increased openness surrounding mental health problems have lowered the threshold for self-reporting ADS. However, this hypothesis lacks support from experimental studies [49] or studies of measurement invariance [50]. Instead, similar increases are observed in diagnostic interviews [51], data on healthcare use [52] and self-harm rates [53]. While increased openness may have impacted on these observed trends as well, the total picture across data sources suggests that factors beyond changes in reporting behavior are at play. The Norwegian population has undergone demographic changes over the observation period, for instance with more immigrants [54]who may face greater life challenges. While some argue that this could partly explain the findings, it does not explain why the observed increase is limited to young adults. Societal conditions for young adults have also changed over the observation period. This generation has grown up in a more performance-driven society, with increasing self-oriented perfectionism [55], less free play [56, 57] and more time spent at home [58], increasing loneliness [59], rising social media use [58, 60], and reported reductions in sleep [61]. Moreover, young adults may increasingly be exposed to anxiety-provoking world events with 24/7 access to media [2]. It seems unlikely that the observed changes are due to a single cause. Rather, the changes should be understood within the framework of a complex adaptive system [62], where multiple interacting factors continuously shape the observed trends.

The decline in depressive symptoms among older adults may reflect that cohorts of older Norwegians live longer without functional impairment, reflecting improvements in health over time [63]. In addition, declining response rates may have increased the selection of healthier participants, particularly following the transition to web-based data collection, which may have amplified selection bias among older adults.

Trends in ADS are consistent across studies for younger age groups, though prevalences differ, particularly among young women. Some of this heterogeneity may reflect methodological factors, such as measurement choice or mode of data collection. For example, lower prevalence is observed in the survey assessing depressive symptoms (HUNT) and in the telephone interview survey. Differences may also reflect variation between samples, underscoring the need for future research to identify subgroups of young adults at increased risk. For instance, Sami women report higher levels of mental health problems compared to non-Sami women [64]. Moreover, migration may be a risk factor for mental health problems [65].

### Strength and limitations

A major strength of the current study is the inclusion of all Norwegian population-based repeated cross-sectional studies, providing the best available evidence base for policy makers. In addition, the replication of similar trends across multiple independent datasets strengthens the robustness of the findings and reduces the likelihood that the observed patterns are artefacts of any single study. However, some limitations should be acknowledged. First, this paper displays data from symptom scales and not diagnostic instruments. There is a global lack of studies using structured diagnostic interviews in population-based samples [66], and symptoms scales are the only measures available in Norway with consistent measure for the past three decades. Thus, the current paper presents the most comprehensive knowledge base on trends in ADS within the Norwegian population over time. Second, to harmonize data, we used the abbreviated HSCL-5, even though the HSCL-25 was available for some studies. While reducing the number of items in the questionnaire may result in some loss of information, research shows that the shorter versions perform very similarly to the full version [27]. Reducing the number of items also increases the prevalence [30], but it is less likely to affect the observed trends. A third limitation is that we investigate changes in internalizing symptoms only, while externalizing symptoms are more frequent among males. Future studies should monitor trends in both internalizing and externalizing difficulties to better understand mental health trends and sex differences. Fourth, participation rates in all surveys decreased over time, and we have limited access to data on non-responders. Non-responders often report worse health outcomes [67]; however, response may also depend on the perceived relevance of the survey topic [68]. While this may affect the reported levels of ADS, its potential impact on observed trends over time is less clear. Finally, while the questionnaires used to measure mental health have remained stable, the mode of data collection changed for some surveys during the study period. Shifts from self-administered paper-and-pencil or online surveys to interviewer-based telephone surveys introduce challenges in interpreting changes over time, as interview modes have been found to increase social desirability and lead to better reports of mental health [34]. However, this issue was addressed by splitting the surveys and avoiding comparisons between time points with different data collection modes. Despite these limitations, our results provide a strong evidence base for the divergent trends in ADS within the adult population from 1995 until 2024.

### Implications and future research

The concerning increase in ADS among young adults remains poorly understood. Although most studies included in the present analysis assess a combination of anxiety and depressive symptoms, findings from the HUNT Study suggest that the rise among young adults is particularly pronounced for anxiety symptoms. Future research should therefore assess symptoms of anxiety and depression separately, using validated instruments, to clarify which specific mental health problems are increasing. Moreover, studies use a wide range of instruments to assess symptoms of anxiety and depression. Understanding changes in ADS requires the ability to compare results across studies. Agreement on common measurements is therefore essential to improve the precision of national trend estimates, as recently emphasized by the International Alliance of Mental Health Research Funders [69].

Our results suggest that that the increase in ADS is mainly concerning young adults. Early adulthood is a critical period for the onset of mental health problems and early onset strongly predicts the risk of future mental health problems, chronicity and impaired functioning [70, 71]. Therefore, increasing ADS in this vulnerable group would be expected to have a greater impact on longer-term mental health and functional outcomes compared to increases in ADS in older age groups. Fortunately, while symptoms of ADS increase the likelihood of developing disorders, treating subclinical symptoms has been shown to the reduce the risk [72]. Furthermore, in a society with an aging population, young adults are essential for sustaining the financial and social framework. Consequently, preventing mental health problems among young adults represents a critical investment in public health [73], especially given the availability of low-threshold interventions [74]. To be able to target preventive efforts, future research should identify sub-groups of young adults who are at increased risk.

### Conclusion

Results from eight population-based studies reveal a clear increase in anxiety and depressive symptoms among young adults in Norway, particularly among young women in the last decade. Our findings indicate that the increase is not limited to emerging adults, as rising levels are also observed among men and women up to their late thirties. While the causes of these changes remain poorly understood, it is evident that they represent an important public health concern. Among adults in midlife, trends in ADS appear more mixed, with most studies showing stability or minor fluctuations. Meanwhile, the mental health of the older adults seems to have either improved or remained stable.

## Data Availability

Data are available upon reasonable request
to the respective data owners.

https://doi.org/10.18712/NSD-NSD0415-V5

https://doi.org/10.18712/NSD-NSD0842-V6

https://doi.org/10.18712/NSD-NSD1327-V9

https://doi.org/10.18712/NSD-NSD2034-V4

https://doi.org/10.18712/NSD-NSD2365-1-V5

https://doi.org/10.18712/NSD-NSD2900-2-V5

https://doi.org/10.18712/NSD-NSD2935-V3

https://doi.org/10.18712/NSD-NSD2995-V2

https://doi.org/10.18712/NSD-NSD3106-V1

https://doi.org/10.18712/NSD-NSD3173-V1

## Declaration of AI assisted copy editing in the writing process

In writing this paper the authors used ChatGPT 4o in order improve readability and language of parts of the written text. After using this tool, the authors reviewed and edited the content as needed and take full responsibility for the content of the publication.

## Statements and Declarations

### Author Contributions

Benedicte Kirkøen and Anne Reneflot contributed to the study conception. All authors contributed to the study design. Astrid MA Eriksen, Ann Ragnhild Broderstad, Anne Høye, Jonas Johansson, Børge Sivertsen, Elin Skretting Lunde, Steinar Krokstad, Thomas Sevenius Nilsen and Marit Knapstad contributed to data collection. Material preparation and analysis were performed by Benedicte Kirkøen, Erik R. Sund and Marita Melhus. The first draft of the manuscript was written by Benedicte Kirkøen and all authors commented on previous versions of the manuscript and revised it critically for important intellectual content. All authors read and approved the final manuscript.

### Ethics approval and consent to participate

We made use of secondary anonymised datasets collected as part of data-collections in the studies HUNT, the Tromsø Study, SAMINOR, FHUS and SHOT or as part of collection of official statistics by Statistics Norway. Because the datasets were anonymised, specific ethical approval was not required for this paper and was waved by the Institutes Data Protection Officer.

