## Supplementary material for "Trends in symptoms of anxiety and depression among adults in Norway: Evidence from eight population-based surveys (1995-2024)": Table 1

**Table 1.** Characteristics of the included population-based cross-sectional studies.

| **Study** | **Year** | **Sample n*** | **Women (%)** | **Outcome** | **Outcome range** | **Mode** | **Population** | **Response rate** |
| --- | --- | --- | --- | --- | --- | --- | --- | --- |
| **Living Conditions Survey** (health and European health interview survey (EHIS))  Statistics Norway | 1998 | 6886 | 53% | HSCL-25 | 1-4 | Paper | National | **72%** |
|  | 2002 | 5313 | 51% | HSCL-25 | 1-4 | Paper | National | **64%** |
|  | 2005 | 4705 | 52% | HSCL-25 | 1-4 | Paper | National | **57%** |
|  | 2008 | 4392 | 53% | HSCL-25 | 1-4 | Paper | National | **50%** |
|  | 2012 | 3895 | 53 % | HSCL-25 | 1-4 | Paper or web | National | **71%** |
|  | 2015 | 8104 | 49.7% | HSCL-5 | 1-4 | Telephone | National | **59%** |
|  | 2019 | 7897 | 49.8% | HSCL-5 | 1-4 | Telephone | National | **57%** |
| **The Trøndelag Health Study (HUNT)** | 1995-1997 | 62 444 | 52.8% | HADS | 0-21 | Paper | Trøndelag County, 20 years + | **70%** |
|  | 2006-2008 | 48 362 | 54.4% | HADS | 0-21 | Paper | Trøndelag County, 20 years + | **54%** |
|  | 2017-2019 | 52 442 | 54.3% | HADS | 0-21 | Paper (online option) | Trøndelag County, 20 years + | **54%** |
| **Quality of Life Survey**  Statistics Norway | 2020** | 17 529 | 51.4% | HSCL-5 | 1-4 | Web | National | **44%** |
|  | 2021 | 17 544 | 51.9% | HSCL-5 | 1-4 | Web | National | **44%** |
|  | 2022 | 15 133 | 51.7% | HSCL-5 | 1-4 | Web | National | **38%** |
|  | 2023 | 17 972 | 51.6 % | HSCL-5 | 1-4 | Web | National | **45%** |
|  | 2024 | 17 229 | 51.6 % | HSCL-5 | 1-4 | Web | National | **43%** |
| **Student’s Health and Wellbeing Study (SHoT)** | 2010 | 5971 | 65% | HSCL-25 | 1-4 | Web | Random sample of full-time students | **23%** |
|  | 2014 | 13 523 | 66.5% | HSCL-25 | 1-4 | Web | Random sample of full-time students | **29%** |
|  | 2018 | 49 754 | 69% | HSCL-25 | 1-4 | Web | All Norwegian full-time students | **31%** |
|  | 2021 | 59 028 | 65.6% | HSCL-5 | 1-4 | Web | All Norwegian full-time students | **34%** |
|  | 2022 | 52 959 | 66.6% | HSCL-25 | 1-4 | Web | All Norwegian full-time students | **35%** |
| **The Tromsø Study** | 2001 | 7021 | 56,7% | HSCL-10 | 1-4 | Paper | Tromsø municipality, 30 years + | **78%** |
|  | 2007-2008 | 12 293 | 53,4% | HSCL-10 | 1-4 | Paper | Tromsø municipality, 30 years + | **66%** |
|  | 2015-2016 | 20 422 | 52,5% | HSCL-10 | 1-4 | Paper and web | Tromsø municipality, 40 years + | **65%** |
| **The Population-based Study on Health and Living Conditions in Regions with Sami and Norwegian Populations**  **(SAMINOR)** | 2003-2004 | 11 304 | 50.7% | HSCL-10 | 1-4 | Paper | 24 municipalities in the North of Norway. 40-69 years | **64%** |
|  | 2012 | 6776 | 52.50% | HSCL-10 | 1-4 | Paper (online option) | 24*** municipalities in the North of Norway. 40-69 years | **32%** |
| **County Public Health Surveys (FHUS) Oslo** | 2020 | 8865 | 56,5% | HSCL-5 | 1-4 | Web | Oslo county | **40%** |
|  | 2021 | 7506 | 55% | HSCL-5 | 1-4 | Web | Oslo county | **35%** |
|  | 2024 | 43 848 | 56% | HSCL-5 | 1-4 | Web | Oslo county | **28%** |
| **County Public Health Surveys (FHUS) Agder** | 2019 | 28 000 | 53,2% | HSCL-5 | 1-4 | Web | Agder county | **46%** |
|  | 2023 | 18 379 | 55,7% | HSCL-5 | 1-4 | Web | Agder county | **32%** |

*Number of participants who completed the measures of symptoms of anxiety and depression (HSCL og HADS)

** The data collection for the survey was conducted from March 9 to March 29, 2020. This means that interviews were carried out both before and after the extensive measures to limit the spread of the coronavirus, which were implemented on March 12. Approximately 25% of the sample responded before March 12, while the remaining 75% of responses were given after March 12. ( <https://www.ssb.no/sosiale-forhold-og-kriminalitet/artikler-og-publikasjoner/_attachment/433414?_ts=17554096418> )

*** 24 municipalities were invited to SAMINOR 1. For SAMINOR 2, the same municipalities were invited, but also the additional municipality Sør-varanger. To make data from the two survey-waves comparable we excluded Sør-varanger from SAMINOR 2, making the invited municipalities in the two waves the same
