## Supplementtable S1 for "Trends in symptoms of anxiety and depression among adults in Norway: Evidence from eight population-based surveys (1995-2024)"

**Table S1.** Response rate for different age groups

| **Study** | **Year** | **Sample n*** | **Population** | **Response rate** |
| --- | --- | --- | --- | --- |
| **Living Conditions Survey.** Statistics Norway | 1998 | 6886 | National | **72%**  By age: not available |
|  | 2002 | 5313 | National | **64%**  By age:  16-24 years: 57%  25-44 year: 63%  45-66 years: 71%  67-79 year: 64% |
|  | 2005 | 4705 | National | **57%**  By age:  16-24 years:49%  25-44 year: 54%  45-66 years: 62%  67-79 year: 58% |
|  | 2008 | 4392 | National | **50%**  By age:  16-24 years: 37%  25-44 year: 45 %  45-66 years: 59%  67-79 year: 57% |
|  | 2012 | 3895 | National | **71%**  By age: not available |
|  | 2015 | 8104 | National | **59%**  By age:  16-24 years: 62%  25-44 year: 55%  45-66 years: 63%  67-79 year: 63%  80+ : 50% |
|  | 2019 | 7897 | National | **57%**  By age:  16-24 years: 55%  25-44 year: 54%  45-66 years: 61%  67-79 year: 62%  80+ : 48% |
| **Study** | **Year** | **Sample n*** | **Population** | **Response rate** |
| **The Trøndelag Health Study (HUNT)** | 1995-1997 | 62 444 | Trøndelag County, 20 years + | **70%**  By age(1)  (women-men)  19-29: 56-42%  30-39: 75-62%  40-49: 82-72%  50-59: 86-77%  60-69: 87-84%  70-79: 79-80%  80-89: 65-68% |
|  | 2006-2008 | 48 362 | Trøndelag County, 20 years + | **54%**  By age (2)  (women-men)  19-29: 38-26%  30-39: 51-36%  40-49: 64-49%  50-59: 71-61%  60-69: 74-66%  70-79: 67-66%  80-89: 39-45% |
|  | 2017-2019 | 52442 | Trøndelag County, 20 years + | **54%**  By age(3)  (women-men)  19-29: 43-31%  30-39: 53-38%  40-49: 59-45%  50-59: 66-54%  60-69: 70-64%  70-79: 68-65%  80-89: 54-55% |
| **Study** | **Year** | **Sample n*** | **Population** | **Response rate** |
| **Quality of life survey.**  Statistics Norway | 2020** | 17.529 | National | **44%**  By age:  18-24 years: 39,8%  25-44 year: 40,3%  45-66 years: 51,7%  67-79 year: 44,8%  80+ : 17,4% |
|  | 2021 | 17.544 | National | **44%**  By age:  18-24 years: 36,7%  25-44 year: 39,4%  45-66 years: 52,3%  67-79 year:48,1%  80+ : 19,1% |
|  | 2022 | 15.133 | National | **38%**  By age  18-24 years: 31%  25-44 year: 33,6%  45-66 years: 45,2%  67-79 year: 41,9%  80+ : 19% |
|  | 2023 | 17.972 | National | **45%**  By age  18-24 years: 40,1%  25-44 year: 43,1%  45-66 years: 52,3%  67-79 year:45,4%  80+ : 19,7% |
|  | 2024 | 17.229 | National | **43%**  By age  18-24 years: 38,1%  25-44 year: 40,7%  45-66 years: 50,7%  67-79 year:44,5%  80+ :21,2% |
| **Study** | **Year** | **Sample n*** | **Population** | **Response rate** |
| **Student’s Health and Wellbeing Study (SHoT)** | 2010 | 5 971 | Random sample of full-time students | **23%** |
|  | 2014 | 13523 | Random sample of full-time students | **29%** |
|  | 2018 | 49754 | Inviting all Norwegian full-time students | **31%** |
|  | 2021 | 59028 | Inviting all Norwegian full-time students | **34%** |
|  | 2022 | 52959 | Inviting all Norwegian full-time students | **35%** |
| **The Tromsø Study** | 2001 | 7021 | Regional. Tromsø municipality, 30+ | **78%**  By age:  (women-men)  30-34: 53-38%  35-39: 86-83%  40-49: 70-61%  50-59: 94-92%  60-69: 91-90%  70-79: 83-87%  80-87: 65-70% |
|  | 2007-2008 | 12 293 | Regional. Tromsø municipality, 30+ | **66%**  By age (4)  (women-men)  30-39: 54-38%  40-49: 65-56%  50-59: 75-66%  60-69: 80-74%  70-79: 66-69%  80-87: 37-36% |
|  | 2015-2016 | 20 422 | Regional. Tromsø municipality, 40+ | **65%**  By age(5)  (women-men)  40-49:65-55%  50-59: 72-65%  60-69: 75-71%  70-79: 68-69%  80-89: 40-51% |
| **Study** | **Year** | **Sample n*** | **Population** | **Response rate** |
| **SAMINOR** | 2003-2004 | 11304 | 24 municipalities in the North of Norway:  40-69 years | **64%**  By age  40-44:57%  45-49:61%  50-54:65%  55-59:66%  60-64:68%  65-69:66% |
|  | 2012 | 6776 | 24*** municipalities in the North of Norway:  40-69 years | **32%**  By age  40-49:28%  50-59: 33%  60-69: 35% |
| **FHUS Oslo** | 2020 | 8 865 | Oslo | **40%**  By age  Women -men  18-29: 32-20%  30-39: 41-29%  40-49: 50-39%  50-59: 56-46%  60-69: 63-52%  70-79: 61-59 %  80+: 27-43% |
|  | 2021 | 7 506 | Oslo | **35%**  By age  Women -men  18-29: 21-13%  30-39: 33-25%  40-49: 43-34%  50-59: 52-43%  60-69: 57-53%  70-79: 55-57 %  80+: 28-47% |
|  | 2024 | 43 848 | Oslo | **28%**  By age  Women- men  18-29: 24-15%  30-39: 29-21%  40-49: 31-24%  50-59: 39-31%  60-69: 44-36%  70-79: 50-47%  80+ 32-25% |
| **FHUS Agder** | 2019 | 28 000 | Agder | **46%**  By age  Women- men  18-29: 44-28%  30-39: 46-33%  40-49: 51-42%  50-59: 56-47%  60-69: 58-59%  70+: 47-56% |
|  | 2023 | 18 379 | Agder | **32%**  By age  Women- men  18-29: 25-12%  30-39: 33-20%  40-49: 37-26%  50-59: 44-32%  60-69: 46-41%  70-79: 43-48%  80+: 35-34% |
