## Supplementary material for "Trends in symptoms of anxiety and depression among adults in Norway: Evidence from eight population-based surveys (1995-2024)"

**Table 1a.** Mean score and percentage scoring above cut-off level for mental health problems by age group for women in the Living Conditions Survey (paper version).

| **Living Conditions Survey (paper version)** | | | | | | | | |
| --- | --- | --- | --- | --- | --- | --- | --- | --- |
| **Year** | **N=** | **m=** | **sd=** | **Reg. Coefficient comparing this year to the year prior** | **P=** | **Percentage scoring above ≥1.80** | **OR for being above cut of in this year compared to the year prior** | **P=** |
| **Women, 20-29 years** | | | | | | | | |
| 1998 | 613 | 1,37 | 0,55 |  |  | 17,94 |  |  |
| 2002 | 393 | 1,33 | 0,48 | -0.01 | 0.46 | 16,79 | 0.99 | 0.88 |
| 2005 | 370 | 1,39 | 0,57 | 0.03 | 0.11 | 18,92 | 1.07 | 0.47 |
| 2008 | 259 | 1,40 | 0,50 | 0.01 | 0.74 | 18,92 | 1.02 | 0.86 |
| 2012 | 238 | 1,55 | 0,64 | 0.06 | **0.01** | 29,83 | 1.37 | **<.01** |
| Age (1998-2012) |  |  |  | -0.02 | **<.01** |  | 0.93 | **<.01** |
| 2012 vs 1998 |  |  |  | 0.06 | **<.01** |  | 1.30 | **<.01** |
| **Women, 30-39 years** | | | | | | | | |
| 1998 | 725 | 1.35 | 0,52 |  |  | 18,48 |  |  |
| 2002 | 536 | 1.34 | 0,54 | -0.004 | 0.74 | 18,10 | 0.99 | 0.87 |
| 2005 | 509 | 1.32 | 0,50 | -0.01 | 0.52 | 17,29 | 0.98 | 0.76 |
| 2008 | 397 | 1.35 | 0,47 | 0.01 | 0.49 | 18,14 | 1.03 | 0.70 |
| 2012 | 273 | 1.39 | 0,55 | 0.02 | 0.32 | 22,71 | 1.13 | 0.19 |
| Age (1998-2012) |  |  |  | 0.003 | 0.38 |  | 1.01 | 0.43 |
| 2012 vs 1998 |  |  |  | 0.01 | 0.66 |  | 1.09 | 0.33 |
| **Women, 40-49 years** | | | | | | | | |
| 1998 | 658 | 1.36 | 0.50 |  |  | 19,00 |  |  |
| 2002 | 527 | 1.34 | 0.50 | -0.009 | 0.50 | 18,22 | 0.97 | 0.67 |
| 2005 | 494 | 1.32 | 0.48 | -0.01 | 0.47 | 17,21 | 0.97 | 0.69 |
| 2008 | 443 | 1.37 | 0.53 | 0.03 | 0.10 | 17,38 | 1.01 | 0.92 |
| 2012 | 335 | 1.37 | 0.52 | -0.003 | 0.83 | 17,91 | 1.02 | 0.82 |
| Age (1998-2012) |  |  |  | -0.002 | 0.48 |  | 1.01 | 0.51 |
| 2012 vs 1998 |  |  |  | 0.001 | 0.96 |  | 0.95 | 0.56 |
| **Women, 50-59 years** | | | | | | | | |
| 1998 | 541 | 1.41 | 0.58 |  |  | 22,00 |  |  |
| 2002 | 492 | 1.36 | 0.54 | -0.02 | 0.15 | 17,48 | 0.87 | 0.07 |
| 2005 | 436 | 1.35 | 0.52 | -0.003 | 0.85 | 19,72 | 1.08 | 0.37 |
| 2008 | 449 | 1.37 | 0.51 | 0.01 | 0.70 | 20,71 | 1.03 | 0.71 |
| 2012 | 397 | 1.33 | 0.49 | -0.02 | 0.29 | 17,13 | 0.89 | 0.20 |
| Age (1998-2012) |  |  |  | -0.01 | 0.28 |  |  |  |
| 2012 vs 1998 |  |  |  | -0.04 | **0.02** |  | 0.86 | 0.06 |
| **Women, 60-69 years** | | | | | | | | |
| 1998 | 370 | 1.42 | 0.55 |  |  | 23,51 |  |  |
| 2002 | 327 | 1.35 | 0.47 | -0.04 | **.03** | 18,96 | 0.82 | **0.06** |
| 2005 | 342 | 1.24 | 0.39 | -0.05 | **<.01** | 11,11 | 0.75 | **<.01** |
| 2008 | 368 | 1.28 | 0.41 | 0.02 | 0.30 | 13,86 | 1.09 | 0.39 |
| 2012 | 375 | 1.29 | 0.45 | 0.006 | 0.71 | 14,40 | 1.03 | 0.80 |
| Age (1998-2012) |  |  |  |  |  |  |  |  |
| 2012 vs 1998 |  |  |  | -0.06 | **<.01** |  | 0.71 | **<.01** |
| **Women, 70-79 years** | | | | | | | | |
| **Year** | **N=** | **m=** | **sd=** | **Reg. Coefficient comparing this year to the year prior** | **P=** | **Percentage scoring above ≥1.80** | **OR for being above cut of in this year compared to the year prior** | **P=** |
| 1998 | 317 | 1.44 | 0.56 |  |  | 23,34 |  |  |
| 2002 | 218 | 1.42 | 0.55 | -0.02 | 0.35 | 22,02 | 0.92 | 0.44 |
| 2005 | 168 | 1.31 | 0.43 | -0.05 | **0.04** | 18,45 | 0.89 | 0.38 |
| 2008 | 185 | 1.32 | 0.42 | 0.001 | 0.97 | 14,05 | 0.85 | 0.22 |
| 2012 | 239 | 1.32 | 0.43 | -0.01 | 0.74 | 15,48 | 1.02 | 0.87 |
| Age (1998-2012) |  |  |  |  |  |  |  |  |
| 2012 vs 1998 |  |  |  | -0.06 | **<.01** |  | 0.78 | **0.02** |

*The Living Conditions Survey (paper version) also included participants aged 80-89 years old. However, we only include data for participants aged 80-89 for surveys that had close to n>100 for each time point. The Living Conditions Survey (paper version) did not fulfill this criteria

**Table 1b.** Mean score and percentage scoring above cut-off level for mental health problems by age group for men in the Living Conditions Survey (paper version).

| **Living Conditions Survey (paper version)** | | | | | | | | |
| --- | --- | --- | --- | --- | --- | --- | --- | --- |
| **Year** | **N=** | **m=** | **sd=** | **Reg. Coefficient comparing this year to the year prior** | **P=** | **Percentage scoring ≥2.00** | **OR for being above cut of in this year compared to the year prior** | **P=** |
| **Men, 20-29 years** | | | | | | | | |
| 1998 | 486 | 1.35 | 0.53 |  |  | 11.73 |  |  |
| 2002 | 325 | 1.33 | 0.51 | -0.01 | 0.59 | 13.23 | 1.06 | 0.59 |
| 2005 | 284 | 1.29 | 0.50 | -0.02 | 0.42 | 10.21 | 0.86 | 0.25 |
| 2008 | 219 | 1.35 | 0.50 | 0.03 | 0.19 | 9.59 | 0.99 | 0.96 |
| 2012 | 197 | 1.38 | 0.58 | 0.01 | 0.59 | 13.71 | 1.22 | 0.19 |
| Age (1998-2012) |  |  |  | -0.01 | 0.55 |  | 0.99 | 0.93 |
| 2012 vs 1998 |  |  |  | 0.01 | 0.86 |  | 1.06 | 0.63 |
| **Men, 30-39 years** | | | | | | | | |
| 1998 | 680 | 1.28 | 0.45 |  |  | 9.12 |  |  |
| 2002 | 483 | 1.28 | 0.45 | 0.001 | 0.91 | 7.66 | 0.94 | 0.56 |
| 2005 | 406 | 1.28 | 0.46 | -0.00 | 0.99 | 9.61 | 1.13 | 0.28 |
| 2008 | 297 | 1.33 | 0.51 | 0.02 | 0.17 | 12.79 | 1.17 | 0.22 |
| 2012 | 228 | 1.27 | 0.40 | 0.03 | 0.11 | 9.65 | 0.80 | 0.15 |
| Age (1998-2012) |  |  |  | 0.01 | 0.85 |  | 0.97 | 0.21 |
| 2012 vs 1998 |  |  |  | 0.01 | 0.92 |  | 1.05 | 0.67 |
| **Men, 40-49 years** | | | | | | | | |
| **Year** | **N=** | **m=** | **sd=** | **Reg. Coefficient comparing this year to the year prior** | **P=** | **Percentage scoring ≥2.00** | **OR for being above cut of in this year compared to the year prior** | **P=** |
| 1998 | 613 | 1.32 | 0.48 |  |  | 10.60 |  |  |
| 2002 | 544 | 1.27 | 0.47 | -0.03 | 0.08 | 9.01 | 0.91 | 0.35 |
| 2005 | 431 | 1.24 | 0.45 | -0.01 | 0.47 | 7.89 | 0.96 | 0.67 |
| 2008 | 417 | 1.32 | 0.51 | 0.04 | **0.02** | 10.31 | 1.15 | 0.21 |
| 2012 | 312 | 1.32 | 0.48 | -0.01 | 0.75 | 11.86 | 1.06 | 0.64 |
| Age (1998-2012) |  |  |  | 0.01 | 0.49 |  | 1.04 | 0.14 |
| 2012 vs 1998 |  |  |  | -0.01 | 0.50 |  | 1.01 | 0.90 |
| **Men, 50-59 years** | | | | | | | | |
| 1998 | 523 | 1.31 | 0.45 |  |  | 11.28 |  |  |
| 2002 | 490 | 1.29 | 0.47 | -0.01 | 0.53 | 11.22 | 0.99 | 0.97 |
| 2005 | 444 | 1.28 | 0.46 | -0.01 | 0.64 | 9.23 | 0.91 | 0.35 |
| 2008 | 388 | 1.29 | 0.44 | 0.01 | 0.65 | 9.54 | 1.02 | 0.82 |
| 2012 | 381 | 1.32 | 0.47 | 0.02 | 0.36 | 13.39 | 1.22 | 0.09 |
| Age (1998-2012) |  |  |  | 0.001 | 0.84 |  | 0.99 | 0.82 |
| 2012 vs 1998 |  |  |  | 0.002 | 0.89 |  | 1.09 | 0.41 |
| **Men, 60-69 years** | | | | | | | | |
| 1998 | 373 | 1.26 | 0.44 |  |  | 8.85 |  |  |
| 2002 | 335 | 1.26 | 0.42 | -0.003 | 0.83 | 8.06 | 0.93 | 0.60 |
| 2005 | 348 | 1.19 | 0.39 | -0.04 | **0.02** | 5.17 | 0.80 | 0.13 |
| 2008 | 365 | 1.24 | 0.40 | 0.03 | 0.11 | 8.49 | 1.26 | 0.11 |
| 2012 | 377 | 1.26 | 0.45 | 0.01 | 0.37 | 7.16 | 0.93 | 0.59 |
| Age (1998-2012) |  |  |  | -0.01 | 0.04 |  | 0.93 | **0.02** |
| 2012 vs 1998 |  |  |  | 0.0001 | 0.96 |  | 0.88 | 0.37 |
| **Men, 70-79 years** | | | | | | | | |
| 1998 | 270 | 1.27 | 0.40 |  |  | 10.37 |  |  |
| 2002 | 202 | 1.25 | 0.48 | -0.02 | 0.36 | 9.41 | 0.88 | 0.48 |
| 2005 | 193 | 1.17 | 0.33 | -0.04 | 0.06 | 4.66 | 0.71 | 0.07 |
| 2008 | 166 | 1.17 | 0.29 | 0.001 | 0.94 | 4.22 | 1.01 | 0.98 |
| 2012 | 183 | 1.23 | 0.36 | 0.03 | 0.19 | 6.01 | 1.11 | 0.59 |
| Age (1998-2012) |  |  |  | 0.001 | 0.71 |  |  |  |
| 2012 vs 1998 |  |  |  | -0.03 | 0.14 |  | 0.70 | **0.056** |

*The Living Conditions Survey (paper version) also included participants aged 80-89 years old. However, we only include data for participants aged 80-89 for surveys that had close to n>100 for each time point. The Living Conditions Survey (paper version) did not fulfill these criteria

**Table 2a.** Mean score and percentage scoring above cut-off level for mental health problems by age group for women in the Living Conditions Survey (telephone version)

| **Living Conditions Survey (telephone version)** | | | | | | | | |
| --- | --- | --- | --- | --- | --- | --- | --- | --- |
| **Year** | **N=** | **m=** | **sd=** | **Reg. Coefficient comparing this year to the year prior** | **P=** | **Percentage scoring ≥ 1.80** | **OR for being above cut of in this year compared to the year prior** | **P=** |
| **Women, 20-29 years** | | | | | | | | |
| 2015 | 585 | 1.36 | 0.54 |  |  | 17.61 |  |  |
| 2019 | 550 | 1.50 | 0.61 | 0.07 | **<.01** | 25.64 | 1.27 | **<.01** |
| Age |  |  |  | -0.02 | 0.01 |  | 0.93 | 0.01 |
| **Women 30-39 years old** | | | | | | | | |
| 2015 | 561 | 1.30 | 0.48 |  |  | 15.15 |  |  |
| 2019 | 621 | 1.40 | 0.55 | 0.05 | **<.01** | 20.00 | 1.18 | **0.03** |
| Age |  |  |  | -0.01 | 0.57 |  | 0.99 | 0.71 |
| **Women 40-49 years old** | | | | | | | | |
| 2015 | 714 | 1.29 | 0.47 |  |  | 13.45 |  |  |
| 2019 | 643 | 1.33 | 0.50 | 0.02 | 0.11 | 14.93 | 1.06 | 0.43 |
| Age |  |  |  | 0.02 | 0.11 |  | 1.00 | 0.99 |
| **Women 50-59 years old** | | | | | | | | |
| 2015 | 685 | 1.22 | 0.40 |  |  | 10.66 |  |  |
| 2019 | 675 | 1.35 | 0.54 | 0.06 | **<.01** | 18.07 | 1.36 | **<.01** |
| Age |  |  |  | 0.01 | 0.78 |  | 1.02 | 0.43 |
| **Women 60-69 years old** | | | | | | | | |
| 2015 | 637 | 1.19 | 0.39 |  |  | 9.58 |  |  |
| 2019 | 587 | 1.23 | 0.40 | 0.02 | **0.05** | 11.24 | 1.09 | 0.34 |
| Age |  |  |  | -0.01 | 0.34 |  | 0.98 | 0.64 |
| **Women 70-79 years old** | | | | | | | | |
| 2015 | 341 | 1.17 | 0.35 |  |  | 7.92 |  |  |
| 2019 | 411 | 1.23 | 0.38 | 0.03 | **0.05** | 11.44 | 1.22 | 0.10 |
| Age |  |  |  | -0.01 | 0.81 |  | 0.97 | 0.57 |
| **Women 80-89 years old** | | | | | | | | |
| 2015 | 177 | 1.19 | 0.41 |  |  | 6.21 |  |  |
| 2019 | 183 | 1.26 | 0.42 | 0.04 | 0.09 | 10.93 | 1.35 | 0.12 |
| Age |  |  |  | 0.01 | 0.22 |  | 1.10 | 0.16 |

**Table 2b.** Mean score and percentage scoring above cut-off level for mental health problems by age group for men in the Living Conditions Survey (telephone version)

| **Living Conditions Survey (telephone version)** | | | | | | | | |
| --- | --- | --- | --- | --- | --- | --- | --- | --- |
| **Year** | **N=** | **m=** | **sd=** | **Reg. Coefficient comparing this year to the year prior** | **P=** | **Percentage scoring ≥2.00** | **OR for being above cut of in this year compared to the year prior** | **P=** |
| **Men, 20-29 years** | | | | | | | | |
| 2015 | 600 | 1.23 | 0.39 |  |  | 6.17 |  |  |
| 2019 | 589 | 1.30 | 0.47 | 0.04 | **<.01** | 10.53 | 1.34 | **0.01** |
| Age |  |  |  | -0.01 | .42 |  | 0.97 | 0.39 |
| **Men 30-39 years old** | | | | | | | | |
| 2015 | 586 | 1.22 | 0.44 |  |  | 7.68 |  |  |
| 2019 | 566 | 1.26 | 0.45 | 0.02 | 0.13 | 8.66 | 1.07 | 0.52 |
| Age |  |  |  | 0.01 | 0.15 |  | 1.05 | 0.16 |
| **Men 40-49 years old** | | | | | | | | |
| 2015 | 710 | 1.17 | 0.39 |  |  | 5.07 |  |  |
| 2019 | 654 | 1.22 | 0.42 | 0.03 | **0.01** | 7.19 | 1.20 | 0.12 |
| Age |  |  |  | 0.01 | 0.57 |  | 1.04 | 0.23 |
| **Men 50-59 years old** | | | | | | | | |
| 2015 | 688 | 1.13 | 0.35 |  |  | 3.78 |  |  |
| 2019 | 701 | 1.21 | 0.42 | 0.04 | **<.01** | 5.56 | 1.22 | 0.12 |
| Age |  |  |  | 0.01 | 0.69 |  | 0.99 | 0.98 |
| **Men 60-69 years old** | | | | | | | | |
| 2015 | 687 | 1.11 | 0.29 |  |  | 2.47 |  |  |
| 2019 | 628 | 1.15 | 0.33 | 0.02 | **0.02** | 4.30 | 1.32 | 0.08 |
| Age |  |  |  | -0.01 | 0.03 |  | 0.89 | 0.30 |
| **Men 70-79 years old** | | | | | | | | |
| 2015 | 368 | 1.07 | 0.21 |  |  | 2.17 |  |  |
| 2019 | 432 | 1.11 | 0.25 | 0.02 | **0.03** | 2.55 | 1.08 | 0.74 |
| Age |  |  |  | 0.01 | 0.39 |  | 1.04 | 0.61 |
| **Men 80-89 years old** | | | | | | | | |
| 2015 | 131 | 1.13 | 0.29 |  |  | 3.05 |  |  |
| 2019 | 135 | 1.17 | 0.37 | 0.02 | 0.31 | 5.19 | 1.34 | 0.36 |
| Age |  |  |  | 0.01 | 0.15 |  | 1.13 | 0.28 |

**Table 3a.** Mean score and percentage scoring above cut-off level for mental health problems by age group for women in HUNT

| **HUNT** | | | | | | | | |
| --- | --- | --- | --- | --- | --- | --- | --- | --- |
|  |  | **Anxiety**  **HADS-A mean score** | | | | **Anxiety**  **HADS- A cut off** | | |
| **Year** | **N=** | **m=** | **SD=** | **Reg. Coefficient comparing this year to the year prior** | **P=** | **Percentage scoring ≥8** | **OR for being above cut of in this year compared to the year prior** | **P=** |
| **Women, 20-29 years** | | | | | | | | |
| 1995-97 | 4655 | 4.44 | 3.19 |  |  | 15.45 |  |  |
| 2006-08 | 1796 | 4.84 | 3.34 | 0.40 | **<.01** | 19.10 | 1.25 | **<.01** |
| 2017-19 | 2399 | 6.08 | 3.95 | 0.70 | **<.01** | 32.00 | 1.57 | **<.01** |
| Age (1995 – 2017) |  |  |  | -0.04 | **<.01** |  | 0.99 | 0.70 |
| 1995-17 vs 2017-19 |  |  |  | 0.79 | **<.01** |  | 1.60 | **<.01** |
| **Women, 30-39 years** | | | | | | | | |
| 1995-97 | 5766 | 4.48 | 3.41 |  |  | 17.07 |  |  |
| 2006-08 | 3036 | 4.37 | 3.39 | 0.03 | 0.41 | 17.79 | 1.07 | **.01** |
| 2017-19 | 2733 | 5.45 | 3.67 | 0.52 | **<.01** | 26.67 | 1.31 | **<.01** |
| Age (1995 – 2017) |  |  |  | -0.02 | 0.04 |  | 0.98 | 0.15 |
| 1995-17 vs 2017-19 |  |  |  | 0.42 | **<.01** |  | 1.31 | **<.01** |
| **Women, 40-49 years** | | | | | | | | |
| **Year** | **N=** | **m=** | **sd=** | **Reg. Coefficient comparing this year to the year prior** | **P=** | **Percentage scoring ≥8** | **OR for being above cut of in this year compared to the year prior** | **P=** |
| 1995-97 | 6300 | 4.53 | 3.48 |  |  | 17.89 |  |  |
| 2006-08 | 4393 | 4.36 | 3.54 | -0.06 | 0.06 | 17.05 | 0.98 | 0.58 |
| 2017-19 | 3844 | 4.85 | 3.67 | 0.24 | **<.01** | 22.09 | 1.17 | **<.01** |
| Age (1995 – 2017) |  |  |  | 0.01 | 0.31 |  | 1.01 | 0.15 |
| 1995-17 vs 2017-19 |  |  |  | 0.13 | **<.01** |  | 1.13 | **<.01** |
| **Women, 50-59 years** | | | | | | | | |
| 1995-97 | 4571 | 4.59 | 3.54 |  |  | 18.64 |  |  |
| 2006-08 | 4919 | 4.36 | 3.56 | -0.12 | **<.01** | 18.01 | 0.97 | 0.37 |
| 2017-19 | 4718 | 4.62 | 3.59 | 0.13 | **<.01** | 20.41 | 1.08 | **<.01** |
| Age (1995 – 2017) |  |  |  | 0.01 | 0.40 |  | 1.001 | 0.59 |
| 1995-17 vs 2017-19 |  |  |  | 0.02 | 0.67 |  | 1.05 | **0.03** |
| **Women, 60-69 years** | | | | | | | | |
| 1995-97 | 3229 | 4.52 | 3.6 |  |  | 17.96 |  |  |
| 2006-08 | 4362 | 4.28 | 3.41 | -0.12 | **<.01** | 16.41 | 0.94 | 0.06 |
| 2017-19 | 4838 | 4.39 | 3.48 | 0.05 | 0.14 | 17.86 | 1.05 | 0.07 |
| Age (1995 – 2017) |  |  |  | -0.003 | 0.78 |  | 0.99 | 0.63 |
| 1995-17 vs 2017-19 |  |  |  | -0.05 | 0.21 |  | 1.00 | 0.89 |
| **Women, 70-79 years** | | | | | | | | |
| 1995-97 | 2290 | 4.24 | 3.49 |  |  | 17.16 |  |  |
| 2006-08 | 2475 | 4.41 | 3.35 | 0.08 | 0.09 | 17.21 | 1.00 | 0.94 |
| 2017-19 | 3535 | 4.25 | 3.23 | -0.08 | 0.07 | 16.18 | 0.96 | 0.27 |
| Age (1995 – 2017) |  |  |  | -0.02 | 0.05 |  | 0.99 | 0.81 |
| 1995-17 vs 2017-19 |  |  |  | -0.01 | 0.97 |  | 0.96 | 0.27 |
| **Women, 80+ years** |  |  |  |  |  |  |  |  |
| 1995-97 | 763 | 4.19 | 3.59 |  |  | 18.35 |  |  |
| 2006-08 | 891 | 3.97 | 3.31 | -0.13 | 0.13 | 14.14 | 0.85 | **0.02** |
| 2017-19 | 1111 | 4.54 | 3.34 | 0.30 | **<.01** | 18.27 | 1.17 | **0.01** |
| Age (1995 – 2017) |  |  |  | -0.08 | <.01 |  | 0.95 | <.01 |
| 1995-17 vs 2017-19 |  |  |  | 0.21 | **<.01** |  | 1.02 | 0.71 |

| **HUNT** | | | | | | | | |
| --- | --- | --- | --- | --- | --- | --- | --- | --- |
|  |  | **Depression**  **HADS-D mean score** | | | | **Depression**  **HADS-D cut off** | | |
| **Year** | **N=** | **m=** | **SD=** | **Reg. Coefficient comparing this year to the year prior** | **P=** | **Percentage scoring ≥8** | **OR for being above cut of in this year compared to the year prior** | **P=** |
| **Women, 20-29 years** | | | | | | | | |
| 1995-97 | 4764 | 2.25 | 2.40 |  |  | 4.22 |  |  |
| 2006-08 | 1790 | 2.26 | 2.40 | 0.22 | **<.01** | 4.58 | 1.20 | **<.01** |
| 2017-19 | 2392 | 3.31 | 3.12 | 0.57 | **<.01** | 10.70 | 1.84 | **<.01** |
| Age (1995 – 2017) |  |  |  | 0.02 | **0.03** |  | 1.02 | 0.28 |
| 1995-17 vs 2017-19 |  |  |  | 0.55 | **<.01** |  | 1.66 | **<.01** |
| **Women, 30-39 years** | | | | | | | | |
| 1995-97 | 5914 | 2.73 | 2.76 |  |  | 6.90 |  |  |
| 2006-08 | 3023 | 2.53 | 2.68 | -0.07 | **0.02** | 6.29 | 0.98 | 0.58 |
| 2017-19 | 2732 | 3.04 | 2.95 | 0.26 | **<.01** | 8.93 | 1.22 | **<.01** |
| Age (1995 – 2017) |  |  |  | 0.02 | <.01 |  | 1.03 | <.01 |
| 1995-17 vs 2017-19 |  |  |  | 0.12 | **<.01** |  | 1.13 | **<.01** |
| **Women, 40-49 years** | | | | | | | | |
| **Year** | **N=** | **m=** | **sd=** | **Reg. Coefficient comparing this year to the year prior** | **P=** | **Percentage scoring ≥8** | **OR for being above cut of in this year compared to the year prior** | **P=** |
| 1995-97 | 6691 | 3.22 | 2.95 |  |  | 9.30 |  |  |
| 2006-08 | 4381 | 2.79 | 2.83 | -0.22 | <.01 | 7.69 | 0.91 | <.01 |
| 2017-19 | 3848 | 2.92 | 2.97 | 0.07 | **0.03** | 9.04 | 1.09 | **0.03** |
| Age (1995 – 2017) |  |  |  | 0.04 | <.01 |  | 1.01 | 0.09 |
| 1995-17 vs 2017-19 |  |  |  | -0.18 | **<.01** |  | 0.97 | 0.33 |
| **Women, 50-59 years** | | | | | | | | |
| 1995-97 | 4562 | 3.74 | 3.10 |  |  | 12.33 |  |  |
| 2006-08 | 4922 | 3.18 | 2.91 | -0.29 | **<.01** | 9.02 | 0.82 | **<.01** |
| 2017-19 | 4709 | 2.94 | 2.94 | -0.11 | **<.01** | 8.43 | 0.97 | 0.39 |
| Age (1995 – 2017) |  |  |  | 0.05 | <.01 |  | 1.02 | 0.05 |
| 1995-17 vs 2017-19 |  |  |  | -0.42 | **<.01** |  | 0.80 | **<.01** |
| **Women, 60-69 years** | | | | | | | | |
| 1995-97 | 4019 | 4.08 | 3.16 |  |  | 14.21 |  |  |
| 2006-08 | 4393 | 3.41 | 2.77 | -0.31 | **<.01** | 8.81 | 0.75 | **<.01** |
| 2017-19 | 4865 | 2.99 | 2.76 | -0.23 | **<.01** | 7.42 | 0.91 | **<.01** |
| Age (1995 – 2017) |  |  |  | 0.03 | <.01 |  | 1.02 | 0.13 |
| 1995-17 vs 2017-19 |  |  |  | -0.54 | **<.01** |  | 0.77 | **<.01** |
| **Women, 70-79 years** | | | | | | | | |
| **Year** | **N=** | **m=** | **sd=** | **Reg. Coefficient comparing this year to the year prior** | **P=** | **Percentage scoring ≥8** | **OR for being above cut of in this year compared to the year prior** | **P=** |
| 1995-97 | 3199 | 4.39 | 3.29 |  |  | 17.54 |  |  |
| 2006-08 | 2569 | 4.01 | 2.90 | -0.23 | **<.01** | 12.61 | 0.77 | **<.01** |
| 2017-19 | 3614 | 3.27 | 2.70 | -0.40 | **<.01** | 7.58 | 0.77 | **<.01** |
| Age (1995 – 2017) |  |  |  | 0.05 | <.01 |  | 1.02 | 0.02 |
| 1995-17 vs 2017-19 |  |  |  | -0.55 | **<.01** |  | 0.63 | **<.01** |
| **Women, 80-89 years** | | | | | | | | |
| 1995-97 | 1042 | 4.78 | 3.53 |  |  | 19.96 |  |  |
| 2006-08 | 971 | 4.33 | 3.12 | **-**0.22 | **<.01** | 15.45 | **0.85** | **<.01** |
| 2017-19 | 1181 | 4.38 | 3.09 | -0.07 | 0.92 | 15.41 | 0.98 | 0.67 |
| Age (1995 – 2017) |  |  |  | 0.08 | <.01 |  | 1.06 | <.01 |
| 1995-17 vs 2017-19 |  |  |  | -0.21 | **<.01** |  | 0.84 | **<.01** |

**Table 3b.** Mean score and percentage scoring above cut-off level for mental health problems by age group for men in HUNT

| **HUNT** | | | | | | | | |
| --- | --- | --- | --- | --- | --- | --- | --- | --- |
|  |  | **Anxiety**  **HADS-A mean score** | | | | **Anxiety**  **HADS-A cut off** | | |
| **Year** | **N=** | **m=** | **SD=** | **Reg. Coefficient comparing this year to the year prior** | **P=** | **Percentage scoring ≥8** | **OR for being above cut of in this year compared to the year prior** | **P=** |
| **Men, 20-29 years** | | | | | | | | |
| 1995-97 | 3953 | 4.05 | 2.92 |  |  | 11.92 |  |  |
| 2006-08 | 1015 | 4.09 | 2.93 | 0.17 | **<.01** | 12.02 | 1.12 | **0.02** |
| 2017-19 | 1356 | 4.92 | 3.34 | 0.45 | **<.01** | 19.03 | 1.39 | **<.01** |
| Age (1995 – 2017) |  |  |  | 0.04 | <.01 |  | 1.04 | <.01 |
| 1995-17 vs 2017-19 |  |  |  | 0.39 | **<.01** |  | 1.30 | **<.01** |
| **Men, 30-39 years** | | | | | | | | |
| 1995-97 | 5119 | 4.14 | 3.11 |  |  | 12.93 |  |  |
| 2006-08 | 1847 | 3.85 | 3.00 | -0.05 | 0.24 | 11.37 | 1.01 | 0.96 |
| 2017-19 | 1696 | 4.76 | 3.31 | 0.44 | **<.01** | 18.81 | 1.36 | **<.01** |
| Age (1995 – 2017) |  |  |  | -0.04 | <.01 |  | 0.98 | 0.09 |
| 1995-17 vs 2017-19 |  |  |  | 0.23 | **<.01** |  | 1.21 | **<.01** |
| **Men, 40-49 years** | | | | | | | | |
| **Year** | **N=** | **m=** | **sd=** | **Reg. Coefficient comparing this year to the year prior** | **P=** | **Percentage scoring ≥8** | **OR for being above cut of in this year compared to the year prior** | **P=** |
| 1995-97 | 5989 | 4.07 | 3.22 |  |  | 14.04 |  |  |
| 2006-08 | 3328 | 3.86 | 3.11 | -0.08 | **0.02** | 12.50 | 0.95 | 0.14 |
| 2017-19 | 2506 | 4.30 | 3.30 | 0.22 | **<.01** | 16.52 | 1.18 | **<.01** |
| Age (1995 – 2017) |  |  |  | -0.02 | 0.03 |  | 0.98 | 0.16 |
| 1995-17 vs 2017-19 |  |  |  | 0.08 | **0.04** |  | 1.07 | **0.03** |
| **Men, 50-59 years** | | | | | | | | |
| 1995-97 | 4655 | 3.88 | 3.15 |  |  | 12.50 |  |  |
| 2006-08 | 4294 | 3.66 | 3.18 | -0.11 | **<.01** | 11.74 | 0.97 | 0.36 |
| 2017-19 | 3479 | 3.99 | 3.27 | 0.17 | **<.01** | 15.18 | 1.16 | **<.01** |
| Age (1995 – 2017) |  |  |  | -0.01 | 0.33 |  | 0.99 | 0.35 |
| 1995-17 vs 2017-19 |  |  |  | 0.04 | 0.20 |  | 1.12 | **<.01** |
| **Men, 60-69 years** | | | | | | | | |
| 1995-97 | 3351 | 3.37 | 2.91 |  |  | 9.19 |  |  |
| 2006-08 | 3924 | 3.23 | 2.88 | -0.09 | **0.01** | 8.46 | 0.95 | 0.17 |
| 2017-19 | 4243 | 3.48 | 3.01 | 0.14 | <.01 | 11.03 | 1.17 | **<.01** |
| Age (1995 – 2017) |  |  |  | -0.05 | <.01 |  | 0.96 | <.01 |
| 1995-17 vs 2017-19 |  |  |  | 0.07 | **0.04** |  | 1.13 | **<.01** |
| **Men, 70-79 years** | | | | | | | | |
| 1995-97 | 2321 | 3.32 | 2.92 |  |  | 9.35 |  |  |
| 2006-08 | 2195 | 3.16 | 2.63 | -0.08 | **0.05** | 6.56 | 0.83 | **<.01** |
| 2017-19 | 3251 | 3.14 | 2.80 | -0.02 | 0.65 | 8.40 | 1.12 | **0.03** |
| Age (1995 – 2017) |  |  |  | 0.01 | 0.35 |  | 1.01 | 0.62 |
| 1995-17 vs 2017-19 |  |  |  | -0.08 | **0.03** |  | 0.95 | 0.33 |
| **Men, 80-89 years** |  |  |  |  |  |  |  |  |
| 1995-97 | 538 | 3.19 | 2.97 |  |  | 10.59 |  |  |
| 2006-08 | 703 | 3.20 | 2.78 | -0.01 | 0.93 | 8.82 | 0.90 | 0.29 |
| 2017-19 | 934 | 3.41 | 3.04 | 0.11 | 0.13 | 9.85 | 1.05 | 0.54 |
| Age (1995 – 2017) |  |  |  | 0.01 | 0.97 |  | 1.01 | 0.78 |
| 1995-17 vs 2017-19 |  |  |  | 0.12 | 0.13 |  | 0.97 | 0.75 |

| **HUNT** | | | | | | | | |
| --- | --- | --- | --- | --- | --- | --- | --- | --- |
|  |  | **Depression**  **HADS-D mean score** | | | | **Depression**  **HADS-D cut off** | | |
| **Year** | **N=** | **Mean score=** | **SD=** | **Reg. Coefficient comparing this year to the year prior** | **P=** | **Percentage scoring ≥8** | **OR for being above cut of in this year compared to the year prior** | **P=** |
| **Men, 20-29 years** | | | | | | | | |
| 1995-97 | 3984 | 2.34 | 2.35 |  |  | 3.89 |  |  |
| 2006-08 | 1016 | 2.57 | 2.59 | 0.29 | **<.01** | 5.81 | 1.39 | **<.01** |
| 2017-19 | 1353 | 3.47 | 3.01 | 0.51 | **<.01** | 10.20 | 1.64 | **<.01** |
| Age (1995 – 2017) |  |  |  | 0.03 | <.01 |  | 1.02 | 0.23 |
| 1995-17 vs 2017-19 |  |  |  | 0.53 | **<.01** |  | 1.68 | **<.01** |
| **Men, 30-39 years** | | | | | | | | |
| 1995-97 | 5225 | 2.92 | 2.69 |  |  | 6.85 |  |  |
| 2006-08 | 1839 | 2.88 | 2.70 | 0.05 | 0.15 | 7.29 | 1.09 | **0.05** |
| 2017-19 | 1697 | 3.64 | 3.06 | 0.39 | **<.01** | 11.61 | 1.34 | **<.01** |
| Age (1995 – 2017) |  |  |  | 0.04 | <.01 |  | 1.01 | 0.29 |
| 1995-17 vs 2017-19 |  |  |  | 0.30 | **<.01** |  | 1.31 | **<.01** |
| **Men, 40-49 years** | | | | | | | | |
| **Year** | **N=** | **m=** | **sd=** | **Reg. Coefficient comparing this year to the year prior** | **P=** | **Percentage scoring ≥8** | **OR for being above cut of in this year compared to the year prior** | **P=** |
| 1995-97 | 6209 | 3.58 | 3.03 |  |  | 10.44 |  |  |
| 2006-08 | 3323 | 3.24 | 2.87 | -0.17 | **<.01** | 8.97 | 0.92 | **0.03** |
| 2017-19 | 2499 | 3.42 | 3.02 | 0.10 | **.01** | 10.16 | 1.07 | 0.11 |
| Age (1995 – 2017) |  |  |  | 0.03 | <.01 |  | 1.02 | 0.02 |
| 1995-17 vs 2017-19 |  |  |  | -0.12 | **<.01** |  | 0.96 | 0.30 |
| **Men, 50-59 years** | | | | | | | | |
| 1995-97 | 4976 | 4.10 | 3.17 |  |  | 13.59 |  |  |
| 2006-08 | 4308 | 3.57 | 2.97 | -0.28 | **<.01** | 10.47 | 0.84 | **<.01** |
| 2017-19 | 3499 | 3.25 | 2.91 | -0.13 | **<.01** | 9.40 | 0.96 | 0.29 |
| Age (1995 – 2017) |  |  |  | 0.03 | <.01 |  | 1.01 | 0.32 |
| 1995-17 vs 2017-19 |  |  |  | -0.44 | **<.01** |  | 0.80 | **<.01** |
| **Men, 60-69 years** | | | | | | | | |
| 1995-97 | 3805 | 4.03 | 3.11 |  |  | 13.85 |  |  |
| 2006-08 | 3938 | 3.78 | 2.88 | -0.13 | **<.01** | 11.10 | 0.87 | **<.01** |
| 2017-19 | 4262 | 3.23 | 2.84 | -0.28 | **<.01** | 8.42 | 0.86 | **<.01** |
| Age (1995 – 2017) |  |  |  | -0.01 | 0.27 |  | 0.99 | 0.91 |
| 1995-17 vs 2017-19 |  |  |  | -0.40 | **<.01** |  | 0.75 | **<.01** |
| **Men, 70-79 years** | | | | | | | | |
| 1995-97 | 2857 | 4.36 | 3.21 |  |  | 16.77 |  |  |
| 2006-08 | 2257 | 4.23 | 2.9 | -0.11 | **0.01** | 13.74 | 0.86 | **<.01** |
| 2017-19 | 3306 | 3.6 | 2.86 | -0.33 | **<.01** | 10.53 | 0.85 | **<.01** |
| Age (1995 – 2017) |  |  |  | 0.08 | <.01 |  | 1.04 | <.01 |
| 1995-17 vs 2017-19 |  |  |  | -0.38 | **<.01** |  | 0.77 | **<.01** |
| **Men, 80-89 years** |  |  |  |  |  |  |  |  |
| 1995-97 | 719 | 5.02 | 3.49 |  |  | 23.09 |  |  |
| 2006-08 | 737 | 4.73 | 3.10 | -0.13 | 0.12 | 19.81 | 0.91 | 0.15 |
| 2017-19 | 972 | 4.73 | 3.27 | -0.03 | 0.68 | 18.83 | 0.94 | 0.35 |
| Age (1995 – 2017) |  |  |  | 0.10 | <.01 |  | 1.07 | <.01 |
| 1995-17 vs 2017-19 |  |  |  | -0.15 | 0.06 |  | 0.87 | **0.02** |

**Table 4a.** Mean score and percentage scoring above cut-off level for mental health problems by age group for women in the Quality of life survey

| **Quality of life survey** | | | | | | | | |
| --- | --- | --- | --- | --- | --- | --- | --- | --- |
| **Year** | **N=** | **m=** | **sd=** | **Reg. Coefficient comparing this year to the year prior** | **P=** | **Percentage scoring above ≥1.80** | **OR for being above cut of in this year compared to the year prior** | **P=** |
| **Women, 20-29 years** | | | | | | | | |
| 2020 | 1494 | 1.91 | 0.73 |  |  | 52,74 |  |  |
| 2021 | 1421 | 2.03 | 0.78 | 0.06 | **<.01** | 59,25 | 1.15 | **<.01** |
| 2022 | 1155 | 2.11 | 0.79 | 0.04 | **.01** | 63,03 | 1.07 | .07 |
| 2023 | 1443 | 2.07 | 0.79 | -0.01 | 0.39 | 61,19 | .98 | .59 |
| 2024 | 1338 | 2.07 | 0.76 | -0.01 | 0.81 | 61.96 | 1.01 | 0.80 |
| Age (2023- 2020) |  |  |  | -0.03 | **<.01** |  | 0.95 | <.01 |
| 2024 vs 2020 |  |  |  | 0.08 | **<.01** |  | 1.22 | **<.01** |
| **Women, 30-39 years** | | | | | | | | |
| 2020 | 1433 | 1.72 | 0.65 |  |  | 42.64 |  |  |
| 2021 | 1501 | 1.77 | 0.68 | 0.02 | 0.07 | 46.30 | 1.07 | 0.057 |
| 2022 | 1244 | 1.86 | 0.70 | 0.04 | **<.01** | 52.01 | 1.12 | **<.01** |
| 2023 | 1600 | 1.87 | 0.73 | 0.01 | 0.32 | 50.38 | 0.98 | 0.66 |
| 2024 | 1580 | 1.89 | 0.71 | 0.01 | 0.39 | 53.29 | 1.06 | 0.09 |
| Age (2023- 2020) |  |  |  | -0.02 | <.01 |  | 0.97 | <.01 |
| 2024 vs 2020 |  |  |  | 0.09 | **<.01** |  | 1.23 | **<.01** |
| **Women, 40-49 years** | | | | | | | | |
| **Year** | **N=** | **m=** | **sd=** | **Reg. Coefficient comparing this year to the year prior** | **P=** | **Percentage scoring above ≥1.80** | **OR for being above cut of in this year compared to the year prior** | **P=** |
| 2020 | 1641 | 1.60 | 0.60 |  |  | 34.67 |  |  |
| 2021 | 1594 | 1.64 | 0.63 | 0.02 | 0.06 | 38.33 | 1.08 | **0.03** |
| 2022 | 1413 | 1.73 | 0.67 | 0.04 | **<.01** | 43.67 | 1.11 | **<.01** |
| 2023 | 1633 | 1.72 | 0.65 | -0.01 | 0.95 | 42.44 | 0.99 | 0.71 |
| 2024 | 1522 | 1.75 | 0.64 | 0.01 | 0.36 | 44.88 | 1.04 | 0.23 |
| Age (2023- 2020) |  |  |  | -0.01 | <.01 |  | 0.97 | <.01 |
| 2024 vs 2020 |  |  |  | 0.07 | **<.01** |  | 1.23 | **<.01** |
| **Women, 50-59 years** | | | | | | | | |
| 2020 | 1727 | 1.54 | 0.60 |  |  | 31.21 |  |  |
| 2021 | 1824 | 1.55 | 0.59 | 0.01 | 0.62 | 33.44 | 1.05 | 0.19 |
| 2022 | 1523 | 1.64 | 0.63 | 0.04 | **<.01** | 38.61 | 1.12 | **<.01** |
| 2023 | 1749 | 1.64 | 0.64 | 0.01 | 0.58 | 38.02 | 0.99 | 0.95 |
| 2024 | 1764 | 1.65 | 0.64 | 0.01 | 0.59 | 39.06 | 1.02 | 0.50 |
| Age (2023- 2020) |  |  |  | -0.01 | <.01 |  | 0.98 | 0.03 |
| 2024 vs 2020 |  |  |  | 0.05 | **<.01** |  | 1.18 | **<.01** |
| **Women, 60-69 years** | | | | | | | | |
| 2020 | 1381 | 1.48 | 0.54 |  |  | 27.44 |  |  |
| 2021 | 1459 | 1.49 | 0.53 | 0.003 | 0.77 | 29.27 | 1.04 | 0.32 |
| 2022 | 1316 | 1.55 | 0.56 | 0.03 | **<.01** | 34.19 | 1.12 | **<.01** |
| 2023 | 1478 | 1.51 | 0.54 | -0.02 | 0.11 | 29.43 | .90 | **0.01** |
| 2024 | 1420 | 1.51 | 0.56 | 0.01 | 0.88 | 29.79 | 1.01 | 0.85 |
| Age (2023- 2020) |  |  |  | -0.01 | <.01 |  | 0.97 | <.01 |
| 2024 vs 2020 |  |  |  | 0.02 | 0.09 |  | 1.05 | .16 |
| **Women, 70-79 years** | | | | | | | | |
| 2020 | 823 | 1.42 | 0.47 |  |  | 26.00 |  |  |
| 2021 | 851 | 1.49 | 0.52 | 0.03 | **0.01** | 31.26 | 1.14 | **0.02** |
| 2022 | 777 | 1.50 | 0.51 | 0.01 | 0.62 | 30.12 | 0.97 | 0.59 |
| 2023 | 902 | 1.47 | 0.53 | -0.02 | 0.23 | 26.50 | 0.91 | 0.09 |
| 2024 | 820 | 1.45 | 0.49 | -0.01 | 0.46 | 25.98 | 0.99 | 0.87 |
| Age (2023- 2020) |  |  |  | -0.01 | 0.04 |  | 0.98 | 0.13 |
| 2024 vs 2020 |  |  |  | 0.01 | 0.28 |  | 1.01 | 0.97 |
| **Women, 80-89 years** | | | | | | | | |
| 2020 | 174 | 1.46 | 0.56 |  |  | 24.14 |  |  |
| 2021 | 164 | 1.49 | 0.54 | 0.02 | 0.45 | 29.27 | 1.15 | 0.25 |
| 2022 | 168 | 1.52 | 0.53 | 0.01 | 0.63 | 27.38 | 0.95 | 0.71 |
| 2023 | 200 | 1.56 | 0.57 | 9.92 | 0.50 | 30.00 | 1.07 | 0.55 |
| 2024 | 191 | 1.50 | 0.56 | -0.03 | 0.35 | 25.65 | 0.89 | 0.32 |
| Age (2023- 2020) |  |  |  | 0.02 | <.01 |  | 1.06 | 0.05 |
| 2024 vs 2020 |  |  |  | 0.02 | 0.38 |  | 1.04 | 0.72 |

**Table 4b.** Mean score and percentage scoring above cut-off level for mental health problems by age group for men in the Quality of life survey

| **Quality of life survey** | | | | | | | | |
| --- | --- | --- | --- | --- | --- | --- | --- | --- |
| **Year** | **N=** | **m=** | **sd=** | **Reg. Coefficient comparing this year to the year prior** | **P=** | **Percentage scoring ≥2.00** | **OR for being above cut of in this year compared to the year prior** | **P=** |
| **Men, 20-29 years** | | | | | | | | |
| 2020 | 1179 | 1.68 | 0.68 |  |  | 31.13 |  |  |
| 2021 | 1087 | 1.78 | 0.70 | 0.05 | **<.01** | 36.06 | 1.12 | **0.01** |
| 2022 | 924 | 1.88 | 0.74 | 0.05 | **<.01** | 40.69 | 1.09 | **0.04** |
| 2023 | 1238 | 1.86 | 0.72 | -0.01 | 0.82 | 40.06 | 1.01 | 0.85 |
| 2024 | 1076 | 1.82 | 0.69 | -0.02 | 0.13 | 35.97 | 0.91 | **0.03** |
| Age (2024- 2020) |  |  |  | -0.01 | 0.79 |  | 0.99 | 0.47 |
| 2024 vs 2020 |  |  |  | 0.07 | **<.01** |  | 1.12 | **0.01** |
| **Men, 30-39 years** | | | | | | | | |
| 2020 | 1241 | 1.68 | 0.67 |  |  | 29.57 |  |  |
| 2021 | 1204 | 1.70 | 0.70 | 0.01 | 0.32 | 31.89 | 1.06 | 0.20 |
| 2022 | 1049 | 1.74 | 0.70 | 0.02 | 0.20 | 34.32 | 1.05 | 0.25 |
| 2023 | 1407 | 1.75 | 0.70 | 0.01 | 0.50 | 33.40 | 0.98 | 0.80 |
| 2024 | 1283 | 1.77 | 0.69 | 0.01 | 0.56 | 35.78 | 1.05 | 0.22 |
| Age (2024- 2020) |  |  |  | -0.01 | 0.03 |  | 0.99 | 0.35 |
| 2024 vs 2020 |  |  |  | 0.05 | **<.01** |  | 1.15 | **0.01** |
| **Men, 40-49 years** | | | | | | | | |
| **Year** | **N=** | **m=** | **sd=** | **Reg. Coefficient comparing this year to the year prior** | **P=** | **Percentage scoring ≥2.00** | **OR for being above cut of in this year compared to the year prior** | **P=** |
| 2020 | 1546 | 1.54 | 0.61 |  |  | 22.57 |  |  |
| 2021 | 1509 | 1.57 | 0.64 | 0.01 | 0.24 | 24.19 | f | 0.27 |
| 2022 | 1208 | 1.63 | 0.65 | 0.03 | **0.01** | 28.15 | 1.11 | **0.02** |
| 2023 | 1488 | 1.67 | 0.70 | 0.02 | 0.06 | 28.23 | 1.01 | 0.73 |
| 2024 | 1337 | 1.66 | 0.64 | -0.01 | 0.38 | 28.20 | 0.99 | 0.84 |
| Age (2024- 2020) |  |  |  | -0.01 | <.01 |  | 0.96 | <.01 |
| 2024 vs 2020 |  |  |  | 0.06 | **<.01** |  | 1.16 | **<.01** |
| **Men, 50-59 years** | | | | | | | | |
| 2020 | 1739 | 1.46 | 0.57 |  |  | 17.65 |  |  |
| 2021 | 1696 | 1.48 | 0.58 | 0.01 | 0.22 | 20.46 | 1.09 | **0.03** |
| 2022 | 1482 | 1.52 | 0.57 | 0.02 | **0.056** | 21.05 | 1.01 | 0.71 |
| 2023 | 1689 | 1.52 | 0.59 | 0.002 | 0.82 | 21.37 | 1.01 | 0.75 |
| 2024 | 1672 | 1.55 | 0.62 | 0.02 | 0.14 | 23.92 | 1.08 | 0.07 |
| Age (2024- 2020) |  |  |  | -0.01 | <.01 |  | 0.96 | <.01 |
| 2024 vs 2020 |  |  |  | 0.05 | **<.01** |  | 1.21 | **<.01** |
| **Men, 60-69 years** | | | | | | | | |
| 2020 | 1451 | 1.37 | 0.51 |  |  | 13.92 |  |  |
| 2021 | 1521 | 1.38 | 0.52 | 0.01 | 0.55 | 14.00 | 1.01 | 0.97 |
| 2022 | 1317 | 1.41 | 0.52 | 0.02 | 0.11 | 15.41 | 1.05 | 0.28 |
| 2023 | 1449 | 1.42 | 0.54 | 0.01 | 0.61 | 17.60 | 1.09 | 0.11 |
| 2024 | 1517 | 1.44 | 0.53 | 0.01 | 0.28 | 16.94 | 0.98 | 0.75 |
| Age (2024- 2020) |  |  |  | -0.01 | <.01 |  | 0.94 | **<.01** |
| 2024 vs 2020 |  |  |  | 0.04 | **<.01** |  | 1.12 | **0.02** |
| **Men, 70-79 years** | | | | | | | | |
| 2020 | 952 | 1.30 | 0.43 |  |  | 10.40 |  |  |
| 2021 | 1005 | 1.34 | 0.46 | 0.02 | 0.10 | 13.23 | 1.15 | **0.05** |
| 2022 | 920 | 1.34 | 0.42 | 0.01 | 0.77 | 10.87 | 0.89 | 0.11 |
| 2023 | 978 | 1.34 | 0.47 | 0.001 | 0.95 | 12.27 | 1.07 | 0.34 |
| 2024 | 986 | 1.36 | 0.48 | 0.01 | 0.35 | 12.78 | 1.02 | 0.72 |
| Age (2024- 2020) |  |  |  | -0.01 | 0.34 |  | 0.99 | 0.68 |
| 2024 vs 2020 |  |  |  | 0.03 | **<.01** |  | 1.12 | 0.10 |
| **Men, 80-89 years** | | | | | | | | |
| 2020 | 171 | 1.39 | 0.45 |  |  | 14.62 |  |  |
| 2021 | 213 | 1.33 | 0.37 | -0.03 | 0.17 | 10.80 | 0.84 | 0.26 |
| 2022 | 218 | 1.36 | 0.42 | 0.01 | 0.48 | 12.39 | 1.07 | 0.64 |
| 2023 | 212 | 1.36 | 0.45 | 0.01 | 0.91 | 15.09 | 1.12 | 0.41 |
| 2024 | 264 | 1.37 | 0.47 | 0.01 | 0.84 | 12.12 | 0.88 | 0.36 |
| Age (2024- 2020) |  |  |  | 0.01 | 0.66 |  | 1.01 | 0.75 |
| 2024 vs 2020 |  |  |  | -0.01 | 0.80 |  | 0.90 | 0.47 |

**Table 5a.** Mean score and percentage scoring above cut-off level for mental health problems by age group for female students in the Student’s Health and Wellbeing Study (SHoT)

| **SHoT**  **Female student 18-28 years old** | | | | | | | | |
| --- | --- | --- | --- | --- | --- | --- | --- | --- |
| **Year** | **N=** | **m=** | **sd=** | **Reg. Coefficient comparing this year to the year prior** | **P=** | **Percentage scoring ≥2.75*** | **OR for being above cut of in this year compared to the year prior** | **P=** |
| 2010 | 3934 | 1.85 | 0.71 |  |  | 13.42 |  |  |
| 2014 | 8221 | 1.98 | 0.72 | 0.07 | **<.01** | 16.59 | 1.13 | **<.01** |
| 2018 | 33477 | 2.12 | 0.79 | 0.07 | **<.01** | 24.00 | 1.26 | **<.01** |
| 2021 | 36610 | 2.42 | 0.77 | 0.15 | **<.01** | 35.39 | 1.32 | **<.01** |
| 2022 | 32306 | 2.18 | 0.81 | -0.12 | **<.01** | 27.05 | 0.82 | **<.01** |
| 2022 vs 2010 |  |  |  | 0.17 | **<.01** |  | 1.55 | **<.01** |

*Recommended cutt-off levels for female students in the Norwegian student population

* Regression analyses for SHoT are weighted to take the differing n between data-collections into account.

**Table 5b.** Mean score and percentage scoring above cut-off level for mental health problems by age group for male students in the Student’s Health and Wellbeing Study (SHoT)

| **SHoT**  **Male student 18-28 years old** | | | | | | | | |
| --- | --- | --- | --- | --- | --- | --- | --- | --- |
| **Year** | **N=** | **m=** | **sd=** | **Reg. Coefficient comparing this year to the year prior** | **P=** | **Percentage scoring ≥2.25*** | **OR for being above cut of in this year compared to the year prior** | **P=** |
| 2010 | 2037 | 1.57 | 0.63 |  |  | 12.67 |  |  |
| 2014 | 3999 | 1.66 | 0.64 | 0.04 | **<.01** | 15.73 | 1.13 | **<.01** |
| 2018 | 14815 | 1.74 | 0.72 | 0.04 | **<.01** | 20.57 | 1.17 | **<.01** |
| 2021 | 19098 | 2.02 | 0.75 | 0.14 | **<.01** | 32.06 | 1.35 | **<.01** |
| 2022 | 15966 | 1.81 | 0.75 | -0.11 | **<.01** | 23.80 | 0.81 | **<.01** |
| 2022 vs 2010 |  |  |  | -0.12 | **<.01** |  | 1.47 | **<.01** |

*Recommended cutt-off levels for female students in the Norwegian student population

* Regression analyses for SHoT are weighted to take the differing n between data-collections into account.

**Table 6a.** Mean score and percentage scoring above cut-off level for mental health problems by age group for women in the Tromsø study

| **Tromsø Survey** | | | | | | | | |
| --- | --- | --- | --- | --- | --- | --- | --- | --- |
| **Year** | **N=** | **m=** | **sd=** | **Reg. Coefficient comparing this year to the year prior** | **P=** | **Percentage scoring ≥1.85** | **OR for being above cut of in this year compared to the year prior** | **P=** |
| **Women, 30-39 years** | | | | | | | | |
| 2001 | 404 | 1.29 | 0.39 |  |  | 8.91 |  |  |
| 2007-08 | 291 | 1.39 | 0.50 | 0.06 | **<.01** | 13.40 | 1.33 | **0.05** |
| Age (2001-2007-08) |  |  |  | -0.01 | 0.13 |  | 0.97 | 0.46 |
| **Women, 40-49 years** | | | | | | | | |
| 2001 | 733 | 1.32 | 0.43 |  |  | 9.96 |  |  |
| 2007-08 | 1877 | 1.34 | 0.40 | -0.01 | 0.65 | 10.66 | 0.95 | 0.42 |
| 2015-16 | 3304 | 1.39 | 0.45 | 0.03 | **<.01** | 13.80 | 1.16 | **<.01** |
| Age (2001- 2015-16) |  |  |  | 0.01 | 0.16 |  | 1.02 | 0.19 |
| 2001 vs. 2015-16 |  |  |  | 0.04 | **<.01** |  | 1.22 | **<.01** |
| **Women, 50-59 years** | | | | | | | | |
| 2001 | 650 | 1.32 | 0.39 |  |  | 10.62 |  |  |
| 2007-08 | 1249 | 1.35 | 0.42 | 0.01 | 0.19 | 11.29 | 1.02 | 0.83 |
| 2015-16 | 3163 | 1.36 | 0.42 | 0.01 | 0.33 | 11.95 | 1.04 | 0.45 |
| Age (2001- 2015-16) |  |  |  | 0.00 | 0.99 |  | 1.01 | 0.67 |
| 2001 vs. 2015-16 |  |  |  | 0.02 | **0.02** |  | 1.08 | 0.26 |
| **Women, 60-69 years** | | | | | | | | |
| 2001 | 1193 | 1.29 | 0.41 |  |  | 8.80 |  |  |
| 2007-08 | 1946 | 1.33 | 0.41 | 0.02 | **0.02** | 10.53 | 1.09 | 0.15 |
| 2015-16 | 2594 | 1.32 | 0.38 | -0.002 | 0.64 | 9.79 | 0.97 | 0.61 |
| Age (2001- 2015-16) |  |  |  | -0.01 | <.01 |  | 0.95 | <.01 |
| 2001 vs. 2015-16 |  |  |  | 0.01 | 0.11 |  | 1.05 | 0.43 |
| **Women, 70-79 years** | | | | | | | | |
| 2001 | 770 | 1.29 | 0.37 |  |  | 7.92 |  |  |
| 2007-08 | 827 | 1.34 | 0.39 | 0.03 | **<.01** | 10.28 | 1.20 | 0.056 |
| 2015-16 | 1264 | 1.26 | 0.31 | -0.04 | **<.01** | 4.98 | 0.68 | **<.01** |
| Age (2001- 2015-16) |  |  |  | 0.002 | 0.37 |  | 1.01 | 0.64 |
| 2001 vs. 2015-16 |  |  |  | -0.02 | **0.03** |  | 0.78 | **<.01** |
| **Women, 80-89 years** |  |  |  |  |  |  |  |  |
| 2001 | 93 |  |  |  |  |  |  |  |
| 2007-08 | 258 | 1.43 | 0.44 | 0.06 |  | 15.89 | 1.41 |  |
| 2015-16 | 338 | 1.29 | 0.35 | -0.08 | **<.01** | 8.28 | 0.67 | **<.01** |
| Age (2001- 2015-16) |  |  |  | 0.02 | 0.02 |  | 1.10 | 0.05 |

*The Tromsø survey also included participants aged 80-89 years old in 2001. However, we only include data for participants aged 80-89 for surveys that had close to n>100 for each time point. The Tromsø survey in 2001 did not fulfill this criteria, and consequently only data from 2007 and 2015 are included for participants 8-89 years.

**Table 6b.** Mean score and percentage scoring above cut-off level for mental health problems by age group for men in the Tromsø study

| **Tromsø study** | | | | | | | | |
| --- | --- | --- | --- | --- | --- | --- | --- | --- |
| **Year** | **N=** | **m=** | **sd=** | **Reg. Coefficient comparing this year to the year prior** | **P=** | **Percentage scoring ≥1.85** | **OR for being above cut of in this year compared to the year prior** | **P=** |
| **Men, 30-39 years** | | | | | | | | |
| 2001 | 272 | 1.19 | 0.29 |  |  | 2.21 |  |  |
| 2007-08 | 208 | 1.26 | 0.31 | 0.03 | 0.07 | 6.73 | 1.53 | 0.14 |
| Age (2001-2007-08) |  |  |  | 0.01 | 0.58 |  | 1.09 | 0.29 |
| **Men, 40-49 years** | | | | | | | | |
| 2001 | 585 | 1.21 | 0.31 |  |  | 5.30 |  |  |
| 2007-08 | 1644 | 1.25 | 0.36 | 0.01 | 0.49 | 7.24 | 1.03 | 0.71 |
| 2015-16 | 2989 | 1.29 | 0.39 | 0.02 | **<.01** | 8.77 | 1.13 | 0.41 |
| Age (2001- 2015-16) |  |  |  | 0.00 | 0.98 |  | 0.99 | 0.69 |
| 2001 vs. 2015-16 |  |  |  | 0.04 | **<.01** |  | 1.29 | **<.01** |
| **Men, 50-59 years** | | | | | | | | |
| 2001 | 340 | 1.19 | 0.29 |  |  | 4.41 |  |  |
| 2007-08 | 1121 | 1.27 | 0.38 | 0.03 | **0.01** | 7.58 | 1.09 | 0.38 |
| 2015-16 | 2730 | 1.26 | 0.38 | 0.001 | 0.81 | 8.39 | 1.07 | 0.29 |
| Age (2001- 2015-16) |  |  |  | -0.003 | 0.13 |  | 0.96 | 0.09 |
| 2001 vs. 2015-16 |  |  |  | 0.03 | **0.01** |  | 1.24 | **0.03** |
| **Men, 60-69 years** | | | | | | | | |
| 2001 | 1137 | 1.17 | 0.28 |  |  | 3.25 |  |  |
| 2007-08 | 1926 | 1.19 | 0.28 | 0.01 | 0.22 | 4.52 | 1.15 | 0.14 |
| 2015-16 | 2444 | 1.19 | 0.29 | 0.004 | 0.28 | 4.21 | 0.98 | 0.84 |
| Age (2001- 2015-16) |  |  |  | -0.01 | <.01 |  | 0.93 | <.01 |
| 2001 vs. 2015-16 |  |  |  | 0.01 | **0.02** |  | 1.11 | 0.24 |
| **Men, 70-79 years** | | | | | | | | |
| **Year** | **N=** | **m=** | **sd=** | **Reg. Coefficient comparing this year to the year prior** | **P=** | **Percentage scoring ≥1.85** | **OR for being above cut of in this year compared to the year prior** | **P=** |
| 2001 | 749 | 1.18 | 0.28 |  |  | 3.20 |  |  |
| 2007-08 | 779 | 1.21 | 0.31 | 0.02 | **0.03** | 4.75 | 1.26 | 0.09 |
| 2015-16 | 1272 | 1.17 | 0.25 | -0.02 | **<.01** | 2.75 | 0.75 | **0.02** |
| Age (2001- 2015-16) |  |  |  | 0.003 | 0.13 |  | 0.99 | 0.87 |
| 2001 vs. 2015-16 |  |  |  | -0.01 | 0.14 |  | 0.89 | 0.38 |
| **Men, 80-89 years** |  |  |  |  |  |  |  |  |
| 2001 | 95 |  |  |  |  |  |  |  |
| 2007-08 | 167 | 1.22 | 0.31 | 0.01 |  | 5.39 | 1.16 |  |
| 2015-16 | 291 | 1.19 | 0.32 | -0.01 | 0.45 | 4.47 | 0.91 | 0.68 |
| Age (2001- 2015-16) |  |  |  | 4.90 | 0.99 |  | 0.99 | 0.98 |

*The Tromsø survey also included participants aged 80-89 years old in 2001. However, we only include data for participants aged 80-89 for surveys that had close to n>100 for each time point. The Tromsø survey in 2001 did not fulfill this criteria, and consequently only data from 2007 and 2015 are included for participants 8-89 years.

**Table 7a.** Mean score and percentage scoring above cut-off level for mental health problems by age group for women in the SAMINOR study

| **SAMINOR** | | | | | | | | |
| --- | --- | --- | --- | --- | --- | --- | --- | --- |
| **Year** | **N=** | **m=** | **sd=** | **Reg. Coefficient comparing this year to the year prior** | **P=** | **Percentage scoring ≥ 1.85** | **OR for being above cut of in this year compared to the year prior** | **P=** |
| **Women 40-49 years old** | | | | | | | | |
| 2003-04 | 2152 | 1.35 | 0.44 |  |  | 11.66 |  |  |
| 2012 | 1258 | 1.38 | 0.48 | 0.03 | **0.03** | 13.83 | 1.22 | 0.07 |
| Age (2003-2012) |  |  |  | 0.01 | 0.77 |  | 1.00 | 0.93 |
| **Women 50-59 years old** | | | | | | | | |
| 2003-04 | 2218 | 1.36 | 0.46 |  |  | 12.58 |  |  |
| 2012 | 1267 | 1.34 | 0.45 | -0.02 | 0.27 | 11.21 | 0.88 | 0.24 |
| Age (2003-2012) |  |  |  | -0.01 | 0.24 |  | 0.98 | 0.32 |
| **Women 60-69 years old** | | | | | | | | |
| 2003-04 | 1357 | 1.28 | 0.37 |  |  | 7.07 |  |  |
| 2012 | 1035 | 1.28 | 0.38 | 0.01 | 0.88 | 8.50 | 1.21 | 0.22 |
| Age (2003-2021) |  |  |  | -0.01 | <.01 |  | 0.90 | <0.01 |

**Table 7a.** Mean score and percentage scoring above cut-off level for mental health problems by age group for men in the SAMINOR study

| **SAMINOR** | | | | | | | | |
| --- | --- | --- | --- | --- | --- | --- | --- | --- |
| **Year** | **N=** | **m=** | **sd=** | **Reg. Coefficient comparing this year to the year prior** | **P=** | **Percentage scoring ≥1.85** | **OR for being above cut of in this year compared to the year prior** | **P=** |
| **Men 40-49 years old** | | | | | | | | |
| 2003-04 | 1897 | 1.27 | 0.42 |  |  | 8.17 |  |  |
| 2012 | 917 | 1.27 | 0.46 | 0.01 | 0.76 | 9.16 | 1.13 | 0.38 |
| Age (2003-2012) |  |  |  | 0.01 | 0.33 |  | 1.01 | 0.56 |
| **Men 50-59 years old** | | | | | | | | |
| 2003-04 | 2229 | 1.26 | 0.39 |  |  | 7.85 |  |  |
| 2012 | 1118 | 1.27 | 0.41 | 0.01 | 0.49 | 8.14 | 1.04 | 0.78 |
| Age (2003-2012) |  |  |  | 0.01 | 0.44 |  | 1.00 | 0.88 |
| **Men 60-69 years old** | | | | | | | | |
| 2003-04 | 1451 | 1.19 | 0.31 |  |  | 4.55 |  |  |
| 2012 | 1181 | 1.20 | 0.32 | 0.01 | 0.48 | 6.10 | 1.37 | 0.07 |
| Age (2003-2012) |  |  |  | -0.01 | 0.01 |  | 0.97 | 0.28 |

**Table 8a.** Mean score and percentage scoring above cut-off level for mental health problems by age group for women in FHUS Oslo

| **FHUS Oslo** | | | | | | | | |
| --- | --- | --- | --- | --- | --- | --- | --- | --- |
| **Year** | **N=** | **m=** | **sd=** | **Reg. Coefficient comparing this year to the year prior** | **P=** | **Percentage scoring ≥1.80** | **OR for being above cut of in this year compared to the year prior** | **P=** |
| **Women, 20-29 years** | | | | | | | | |
| 2020 | 953 | 2.08 | 0.81 |  |  | 60.76 |  |  |
| 2021 | 530 | 2.08 | 0.80 | 0.01 | 0.89 | 60.94 | 1.01 | 0.80 |
| 2024 | 5 037 | 2.08 | 0.78 | 0.01 | 0.95 | 62.95 | 1.04 | 0.45 |
| Age (2020-2024) |  |  |  | -0.04 | <.01 |  | 0.92 | <.01 |
| 2020 vs. 2024 |  |  |  | 0.01 | 0.77 |  | 1.05 | 0.16 |
| **Women, 30-39 years** | | | | | | | | |
| 2020 | 1099 | 1.87 | 0.73 |  |  | 51.87 |  |  |
| 2021 | 925 | 1.81 | 0.72 | -0.03 | 0.11 | 45.30 | 0.89 | **<0.01** |
| 2024 | 5 179 | 1.93 | 0.74 | 0.06 | **<.01** | 53.89 | 1.19 | **<.01** |
| Age (2020-2024) |  |  |  | -0.01 | 0.01 |  | 0.97 | 0.02 |
| 2020 vs. 2024 |  |  |  | 0.03 | **0.01** |  | 1.04 | 0.18 |
| **Women, 40-49 years** | | | | | | | | |
| 2020 | 920 | 1.71 | 0.67 |  |  | 42.72 |  |  |
| 2021 | 815 | 1.67 | 0.68 | -0.01 | 0.38 | 38.04 | 0.92 | 0.07 |
| 2024 | 4 211 | 1.81 | 0.71 | 0.07 | **<.01** | 46.62 | 1.19 | **<.01** |
| Age (2020-2024) |  |  |  | -0.01 | <.01 |  | 0.96 | <.01 |
| 2020 vs. 2024 |  |  |  | 0.05 | **<.01** |  | 1.08 | **0.05** |
| **Year** | **N=** | **m=** | **sd=** | **Reg. Coefficient comparing this year to the year prior** | **P=** | **Percentage scoring ≥1.80** | **OR for being above cut of in this year compared to the year prior** | **P=** |
| **Women, 50-59 years** | | | | | | | | |
| 2020 | 872 | 1.66 | 0.60 |  |  | 39.56 |  |  |
| 2021 | 812 | 1.58 | 0.60 | -0.04 | **0.02** | 32.51 | 0.87 | <.01 |
| 2024 | 4 061 | 1.69 | 0.65 | 0.06 | **<.01** | 40.63 | 1.19 | <.01 |
| Age (2020-2024) |  |  |  | -0.01 | <.01 |  | 0.97 | 0.03 |
| 2020 vs. 2024 |  |  |  | 0.02 | 0.09 |  | 1.02 | 0.54 |
| **Women, 60-69 years** | | | | | | | | |
| 2020 | 728 | 1.56 | 0.56 |  |  | 32.55 |  |  |
| 2021 | 668 | 1.47 | 0.52 | -0.4 | **<.01** | 27.50 | 0.88 | **0.04** |
| 2024 | 3 133 | 1.52 | 0.58 | 0.03 | **0.02** | 29.17 | 1.04 | 0.36 |
| Age (2020-2024) |  |  |  | -0.01 | 0.19 |  | 0.97 | 0.12 |
| 2020 vs. 2024 |  |  |  | -0.02 | 0.18 |  | 0.93 | 0.09 |
| **Women, 70-79 years** | | | | | | | | |
| 2020 | 322 | 1.57 | 0.55 |  |  | 35.71 |  |  |
| 2021 | 306 | 1.54 | 0.56 | -0.02 | 0.41 | 31.70 | 0.89 | 0.22 |
| 2024 | 1 968 | 1.47 | 0.53 | -0.03 | **0.03** | 24.95 | 0.84 | **0.01** |
| Age (2020-2024) |  |  |  | -0.01 | 0.96 |  | 0.98 | 0.57 |
| 2020 vs. 2024 |  |  |  | -0.05 | **<.01** |  | 0.78 | **<.01** |

* Regression analyses for FHUS Oslo are weighted to take the differing n between data-collections into account.

*The FHUS Oslo also included participants aged 80-89 years old. However, we only include data for participants aged 80-89 for surveys that had close to n>100 for at least two time points. The FHUS Oslo in 2020 and 2021 did not fulfill this criteria, and consequently only data for participants 8-89 years are not included for FHUS Oslo.

**Table 8b.** Mean score and percentage scoring above cut-off level for mental health problems by age group for men in FHUS Oslo

| **FHUS Oslo** | | | | | | | | |
| --- | --- | --- | --- | --- | --- | --- | --- | --- |
| **Year** | **N=** | **m=** | **sd=** | **Reg. Coefficient comparing this year to the year prior** | **P=** | **Percentage scoring ≥2.00** | **OR for being above cut of in this year compared to the year prior** | **P=** |
| **Men, 20-29 years** | | | | | | | | |
| 2020 | 524 | 1.77 | 0.69 |  |  | 34.54 |  |  |
| 2021 | 309 | 1.83 | 0.76 | 0.06 | **0.03** | 36.89 | 1.13 | 0.09 |
| 2024 | 3 007 | 1.92 | 0.75 | 0.04 | 0.06 | 43.40 | 1.14 | **0.04** |
| Age (2020-2024) |  |  |  | -0.02 | **0.01** |  | 0.95 | **0.01** |
| 2020 vs. 2024 |  |  |  | 0.08 | **<.01** |  | 1.20 | **<.01** |
| **Men, 30-39 years** | | | | | | | | |
| 2020 | 846 | 1.67 | 0.69 |  |  | 29.43 |  |  |
| 2021 | 723 | 1.65 | 0.69 | 0.01 | 0.74 | 27.80 | 0.99 | 0.94 |
| 2024 | 3 978 | 1.85 | 0.74 | 0.10 | <.01 | 39.92 | 1.31 | <0.01 |
| Age (2020-2024) |  |  |  | -0.01 | 0.21 |  | 0.97 | 0.02 |
| 2020 vs. 2024 |  |  |  | 0.09 | **<.01** |  | 1.27 | **<.01** |
| **Men, 40-49 years** | | | | | | | | |
| 2020 | 789 | 1.57 | 0.65 |  |  | 24.71 |  |  |
| 2021 | 689 | 1.55 | 0.60 | -0.01 | 0.79 | 21.48 | 0.95 | 0.38 |
| 2024 | 3 423 | 1.77 | 0.73 | 0.11 | **<.01** | 35.26 | 1.41 | **<.01** |
| Age (2020-2024) |  |  |  | -0.02 | <.01 |  | 0.96 | <.01 |
| 2020 vs. 2024 |  |  |  | 0.10 | **<.01** |  | 1.28 | **<.01** |
| **Year** | **N=** | **m=** | **sd=** | **Reg. Coefficient comparing this year to the year prior** | **P=** | **Percentage scoring ≥2.00** | **OR for being above cut of in this year compared to the year prior** | **P=** |
| **Men, 50-59 years** | | | | | | | | |
| 2020 | 753 | 1.50 | 0.61 |  |  | 21.12 |  |  |
| 2021 | 713 | 1.45 | 0.57 | -0.02 | 0.21 | 18.65 | 0.95 | 0.36 |
| 2024 | 3 533 | 1.62 | 0.66 | 0.09 | **<.01** | 28.96 | 1.33 | **<0.01** |
| Age (2020-2024) |  |  |  | -0.01 | 0.04 |  | 0.97 | 0.09 |
| 2020 vs. 2024 |  |  |  | 0.06 | **<.01** |  | 1.23 | **<.01** |
| **Men, 60-69 years** | | | | | | | | |
| 2020 | 547 | 1.39 | 0.52 |  |  | 16.82 |  |  |
| 2021 | 567 | 1.34 | 0.49 | -0.02 | 0.14 | 12.17 | 0.85 | **0.04** |
| 2024 | 2 680 | 1.45 | 0.58 | 0.05 | **<.01** | 18.99 | 1.31 | **<.01** |
| Age (2020-2024) |  |  |  | -0.01 | <.01 |  | 0.94 | 0.01 |
| 2020 vs. 2024 |  |  |  | 0.03 | **0.01** |  | 1.08 | 0.19 |
| **Men, 70-79 years** | | | | | | | | |
| 2020 | 312 | 1.29 | 0.40 |  |  | 9.62 |  |  |
| 2021 | 306 | 1.23 | 0.35 | -0.03 | 0.11 | 6.86 | 0.88 | 0.31 |
| 2024 | 1 879 | 1.34 | 0.47 | 0.05 | **<.01** | 12.56 | 1.41 | **<.01** |
| Age (2020-2024) |  |  |  | -0.01 | 0.78 |  | 1.01 | 0.84 |
| 2020 vs. 2024 |  |  |  | 0.02 | 0.09 |  | 1.16 | 0.16 |

*The FHUS Oslo also included participants aged 80-89 years old. However, we only include data for participants aged 80-89 for surveys that had n>100 for at least two time points. The FHUS Oslo in 2020 and 2021 did not fulfill this criteria, and consequently only data for participants 8-89 years are not included for FHUS Oslo.

**Table 9a.** Mean score and percentage scoring above cut-off level for mental health problems by age group for women in FHUS Agder

| **FHUS Agder** | | | | | | | | |
| --- | --- | --- | --- | --- | --- | --- | --- | --- |
| **Year** | **N=** | **m=** | **sd=** | **Reg. Coefficient comparing this year to the year prior** | **P=** | **Percentage scoring ≥1.80** | **OR for being above cut of in this year compared to the year prior** | **P=** |
| **Women, 20-29 years** | | | | | | | | |
| 2019 | 2 532 | 1.84 | 0.74 |  |  | 48.89 |  |  |
| 2023 | 1 174 | 2.03 | 0.81 | 0.10 | **<.01** | 57.92 | 1.21 | **<.01** |
| Age |  |  |  | -0.04 | <.01 |  | 0.93 | <.01 |
| **Women, 30-39 years** | | | | | | | | |
| 2019 | 2 578 | 1.63 | 0.66 |  |  | 35.80 |  |  |
| 2023 | 1 647 | 1.83 | 0.75 | 0.10 | **<.01** | 48.27 | 1.29 | **<.01** |
| Age |  |  |  | -0.02 | <.01 |  | 0.95 | <.01 |
| **Women, 40-49 years** | | | | | | | | |
| 2019 | 2 994 | 1.53 | 0.58 |  |  | 29.73 |  |  |
| 2023 | 1 847 | 1.68 | 0.69 | 0.08 | **<.01** | 39.52 | 1.24 | **<.01** |
| Age |  |  |  | -0.01 | <.01 |  | 0.96 | <.01 |
| **Women, 50-59 years** | | | | | | | | |
| 2019 | 2 929 | 1.48 | 0.57 |  |  | 26.90 |  |  |
| 2023 | 2 212 | 1.54 | 0.63 | 0.03 | **<.01** | 30.06 | 1.08 | **0.01** |
| Age |  |  |  | -0.01 | 0.50 |  | 0.99 | 0.17 |
| **Women, 60-69 years** | | | | | | | | |
| 2019 | 2 268 | 1.37 | 0.48 |  |  | 19.93 |  |  |
| 2023 | 1 837 | 1.42 | 0.53 | 0.03 | **<.01** | 23.57 | 1.12 | **<.01** |
| Age |  |  |  | -0.02 | <.01 |  | 0.95 | <.01 |
| **Women, 70-79 years** | | | | | | | | |
| 2019 | 911 | 1.38 | 0.46 |  |  | 20.86 |  |  |
| 2023 | 1 110 | 1.36 | 0.46 | -0.01 | 0.20 | 17.75 | 0.90 | 0.08 |
| Age |  |  |  | 0.01 | 0.81 |  | 1.01 | 0.91 |
| **Women, 80-89 years** |  |  |  |  |  |  |  |  |
| 2019 | 106 | 1.34 | 0.43 |  |  | 20.75 |  |  |
| 2023 | 167 | 1.41 | 0.48 | 0.04 | 0.22 | 20.96 | 1.02 | 0.90 |
| Age |  |  |  | -0.01 | 0.37 |  | 0.97 | 0.64 |

**Table 9b.** Mean score and percentage scoring above cut-off level for mental health problems by age group for men in FHUS Agder

| **FHUS Agder** | | | | | | | | |
| --- | --- | --- | --- | --- | --- | --- | --- | --- |
| **Year** | **N=** | **m=** | **sd=** | **Reg. Coefficient comparing this year to the year prior** | **P=** | **Percentage scoring ≥2.00** | **OR for being above cut of in this year compared to the year prior** | **P=** |
| **Men, 20-29 years** | | | | | | | | |
| 2019 | 1 728 | 1.67 | 0.68 |  |  | 29.92 |  |  |
| 2023 | 601 | 1.84 | 0.80 | 0.09 | **<.01** | 38.10 | 1.20 | **<.01** |
| Age |  |  |  | -0.01 | 0.05 |  | 0.98 | 0.31 |
| **Men, 30-39 years** | | | | | | | | |
| 2019 | 1 920 | 1.61 | 0.69 |  |  | 25.21 |  |  |
| 2023 | 1 036 | 1.76 | 0.78 | 0.07 | **<.01** | 33.49 | 1.23 | **<.01** |
| Age |  |  |  | -0.01 | <.01 |  | 0.96 | <.01 |
| **Men, 40-49 years** | | | | | | | | |
| 2019 | 2 547 | 1.50 | 0.61 |  |  | 20.73 |  |  |
| 2023 | 1 360 | 1.62 | 0.69 | 0.06 | **<.01** | 27.21 | 1.20 | **<.01** |
| Age |  |  |  | -0.01 | <.01 |  | 0.98 | 0.13 |
| **Men, 50-59 years** | | | | | | | | |
| 2019 | 2 658 | 1.40 | 0.55 |  |  | 16.67 |  |  |
| 2023 | 1 678 | 1.49 | 0.63 | 0.04 | **<.01** | 20.80 | 1.15 | **<.01** |
| Age |  |  |  | -0.01 | 0.01 |  | 0.97 | 0.02 |
| **Year** | **N=** | **m=** | **sd=** | **Reg. Coefficient comparing this year to the year prior** | **P=** | **Percentage scoring ≥2.00** | **OR for being above cut of in this year compared to the year prior** | **P=** |
| **Men, 60-69 years** | | | | | | | | |
| 2019 | 2 467 | 1.32 | 0.48 |  |  | 12.16 |  |  |
| 2023 | 1 770 | 1.36 | 0.52 | 0.02 | **<.01** | 13.28 | 1.06 | 0.23 |
| Age |  |  |  | -0.01 | <.01 |  | 0.94 | <.01 |
| **Men, 70-79 years** | | | | | | | | |
| 2019 | 1 210 | 1.25 | 0.38 |  |  | 7.85 |  |  |
| 2023 | 1 341 | 1.27 | 0.40 | 0.01 | 0.09 | 7.98 | 1.02 | 0.81 |
| Age |  |  |  | -0.01 | 0.56 |  | 0.96 | 0.23 |
| **Men, 80-89 years** |  |  |  |  |  |  |  |  |
| 2019 | 182 | 1.30 | 0.45 |  |  | 9.89 |  |  |
| 2023 | 226 | 1.30 | 0.43 | -0.01 | 0.92 | 9.73 | 0.99 | 0.99 |
| Age |  |  |  | -0.01 | 0.96 |  | 0.97 | 0.63 |
